# Maternal health factors driving breastfeeding success: an evidence triangulation framework

**DOI:** 10.64898/2026.04.03.26349172

**Authors:** Maeregu Woldeyes Arisido, Carolina Borges, Nancy McBride, Claudia Giambartolomei, Robin Joaquim Hofmeister, Zoltan Kutalik, Maria Christine Magnus, Luisa Zuccolo

## Abstract

**Background:** Despite well-established benefits to mothers and children, breastfeeding rates fall short of WHO recommendations world-wide. Breastfeeding is a highly complex behaviour, affected by known system-level barriers and less well-established individual-level factors. We aimed to inform breastfeeding support by identifying maternal and perinatal determinants of breastfeeding outcomes and modifiable pathways linking education to breastfeeding.

**Methods:** We used evidence triangulation design from complementary genetic and questionnaire-based analyses across four European cohorts including 72,653 mothers and 317,651 offspring. Five breastfeeding outcomes were assessed: initiation, establishment at 2 months, sustained breastfeeding at 6 months, and exclusive and overall breastfeeding duration. Cohort-specific breastfeeding GWAS were meta-analysed and used in two-sample Mendelian randomization (MR) analyses of 17 maternal and perinatal exposures. MR-based mediation assessed pathways linking educational attainment to breastfeeding, and robustness was evaluated using offspring-genotype adjustment, observational association and MR sensitivity analyses.

**Findings:** Genetically predicted higher education, lower body-mass index (BMI), and lower liability to smoking, insomnia and depression were associated with more favorable breastfeeding outcomes. A 1SD (3.4-year) increase in education duration increased initiation odds (OR: 2.32, 95% CI: 1.94,2.77) and prolonged exclusive breastfeeding (β=0.21SD, 95% CI: 0.17,0.24), whereas higher BMI reduced initiation (OR: 0.77, 95% CI: 0.66,0.90). Smoking, depression and BMI mediated 26%, 14% and 12% of education’s effect on exclusive breastfeeding, respectively. There was little evidence for effects of blood pressure, cholesterol or perinatal factors. Findings were consistent across sensitivity analyses.

**Interpretation:** These findings highlight that breastfeeding support could be achieved through a more holistic approach to maternal physical and mental health before and during pregnancy, and that targeted strategies would reduce maternal and infant health inequalities.

**Funding:** This study was supported by Human Technopole’s core funding. Details on cohort-specific funding are provided in Supplementary Material.

**Panel: Research in context:** *Evidence before this study:* Existing evidence on maternal determinants of breastfeeding largely comes from observational studies, which report heterogeneous associations with sociodemographic, cardiometabolic, behavioural and mental health factors. We considered previous observational studies and reviews on breastfeeding initiation, duration and exclusivity, as well as genetic epidemiology studies of pregnancy and reproductive traits. Few studies have used genetically anchored approaches to assess breastfeeding determinants, and no previous MR study has systematically evaluated a broad range of maternal and perinatal exposures across multiple breastfeeding outcomes.

*Added value of this study:* Using data from 72,653 mothers and 317,651 offspring from four European cohorts, we estimated genetic associations with five breastfeeding outcomes and used them in MR analyses of 17 maternal and perinatal exposures. We combined MR, mediation analysis, offspring-genotype adjustment, multivariable regression and sensitivity analyses to triangulate evidence. Higher educational attainment, lower BMI, and lower liability to smoking, insomnia and depression were associated with more favourable breastfeeding outcomes, and BMI, WHR and smoking partly mediated educational differences in breastfeeding.

*Implications of all the available evidence:* These findings suggest that breastfeeding support should be integrated with broader maternal health strategies, including cardiometabolic health, smoking cessation and mental health support from the preconception period onwards. They also highlight opportunities to identify women at higher risk of breastfeeding difficulties after initiation and to test whether interventions targeting these pathways improve breastfeeding continuation and reduce inequalities.

## Introduction

Breastfeeding is essential for maternal and infant health.^1,2^ It supports brain development, protects children from malnutrition, infectious disease and death, and may lower later risk of obesity and chronic disease.^1^ For mothers, prolonged breastfeeding reduces the risk of oncological and cardiovascular disease.^3^ Global breastfeeding rates remain low: 75% of women do not meet exclusive breastfeeding guidelines,^4^ and only 29% of children receive breast milk at 12 months of age.^5^ Alongside strategies addressing systemic barriers, including universal breastfeeding support and parental leave,^2^ identifying factors that impair or foster breastfeeding is crucial.

Breastfeeding is a complex outcome shaped by behavioral and physiological processes and by mother–infant interactions.^4^ A woman’s ability to breastfeed depends, among other factors, on maternal and infant behavioral traits and physical responses to birth and postpartum challenges.^4^ Breastfeeding rates vary widely by sociodemographic context, particularly by educational attainment.^6^ Physical and mental health also likely play key roles, including elevated body mass index (BMI),^7^ smoking,^8^ and type 2 diabetes (T2D),^9^ insomnia,^10^ depression^4^ and post-traumatic stress disorder (PTSD).^11^ Perinatal factors, including gestational duration and birth weight, have also been linked to breastfeeding outcomes.^12,13^

Existing evidence largely comes from observational studies with heterogeneous findings that cannot reliably distinguish causal from confounded associations. Recent cohort evidence also highlights the importance of social disadvantage and maternal mental health in pregnancy and early parenthood.^14^ For education, which is linked to improved breastfeeding, key questions include whether associations reflect sociocultural and behavioral factors influencing the initial choice to breastfeed rather than persistent effects on lactation, and which modifiable factors could serve as public health intervention targets.

Triangulating evidence across genetic and more traditional, questionnaire-based epidemiological approaches has been shown to yield robust results on both drivers^15^ and cardiovascular effects^16^ of pregnancy outcomes and reproductive factors, as also shown on pre-eclampsia.^17^ However, no previous genetically anchored Mendelian Randomization (MR) study has assessed maternal and perinatal drivers of breastfeeding success, due to the lack of genetic associations with breastfeeding outcomes.

Here, we estimate the effects of a broad range of maternal and perinatal factors on breastfeeding success, assess cardiometabolic and psychiatric pathways explaining the role of educational attainment, and evaluate robustness of results through complementary genetic and questionnaire-based associations combining substantial original and publicly-available data.

## Methods

### Study Design and Setting

We used a genetic epidemiological design to assess effects of maternal and perinatal exposures on five breastfeeding outcomes using MR, quantify cardiometabolic and psychiatric mediation of the education–breastfeeding association, and evaluate robustness using MR sensitivity analyses and multivariable regression (Figure 1A–C). Reporting followed the STROBE-MR guideline.^18^

**Figure 1.**
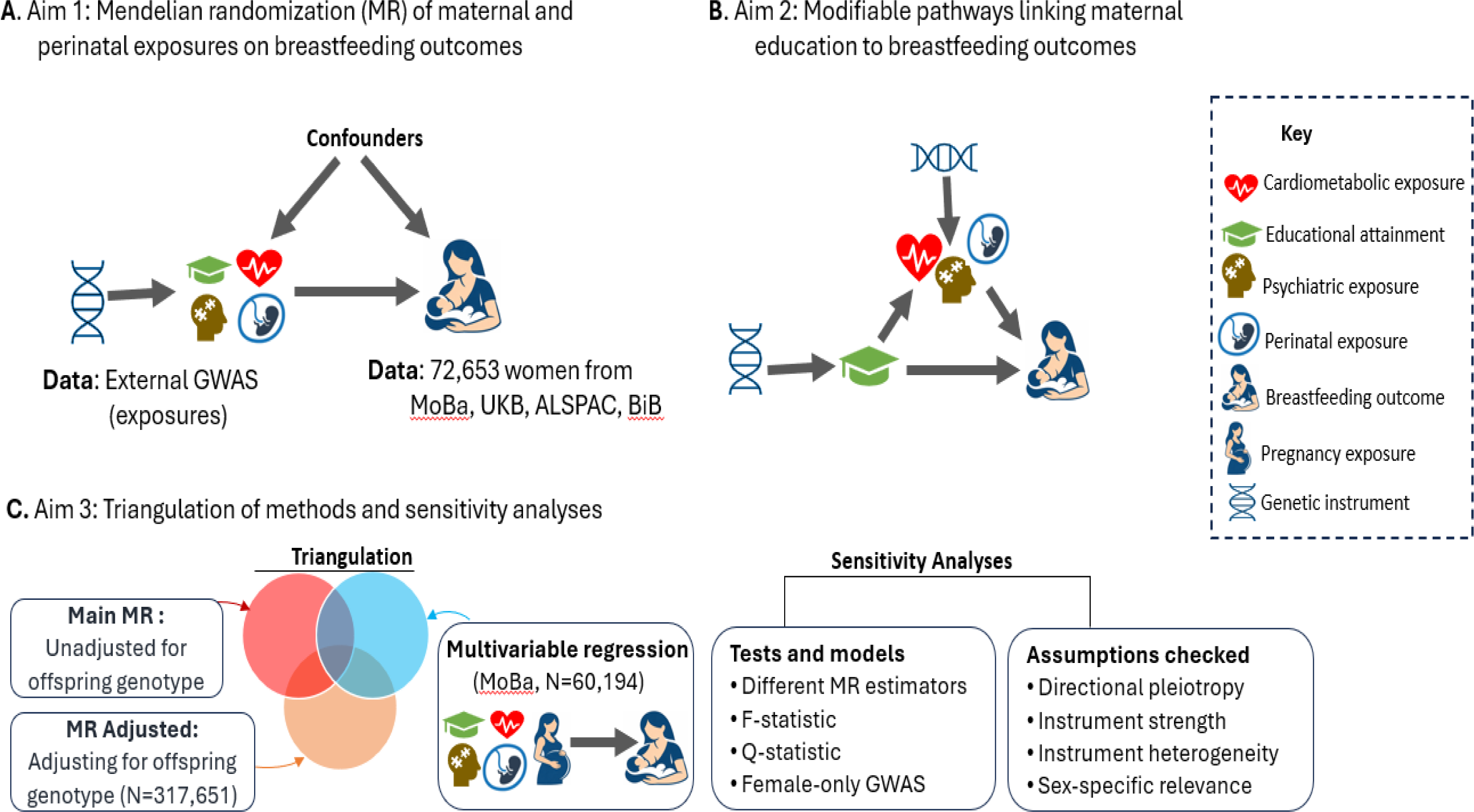
Study design. **(A)** Two-sample Mendelian randomization (MR) estimated effects of maternal exposures on breastfeeding outcomes, using breastfeeding genetic association estimates generated from individual-level data on 72,653 mothers. **(B)** Two-step MR assessed cardiometabolic and psychiatric mediation of the education–breastfeeding association. (**C)** Robustness was assessed through offspring-genotype adjustment, multivariable regression, alternative MR estimators, and tests of instrument strength, heterogeneity and horizontal pleiotropy. ALSPAC, Avon Longitudinal Study of Parents and Children; BiB, Born in Bradford; GWAS, genome-wide association study; MoBa, Norwegian Mother, Father and Child Cohort Study; UKB, UK Biobank.

### Study populations and data sources

We included 72,653 mothers and 317,651 offspring from four European cohorts: the Norwegian Mother, Father and Child Cohort Study (MoBa; N=60,192 mothers), the Avon Longitudinal Study of Parents and Children (ALSPAC; N=6,826), Born in Bradford (BiB; N=1,459) and UK Biobank (UKB; N=4,174; Supplementary Table S1). Women were eligible if they were of European ancestry, had at least one breastfeeding outcome, had a singleton birth, and delivered an infant without a severe known congenital anomaly. In UKB, breastfeeding was reported retrospectively by offspring participants, and maternal–offspring pairs were inferred from genetic relatedness.^19^ Offspring genotype analyses used corresponding offspring samples from the same cohorts. Further recruitment and eligibility details are provided in the Supplementary Material.

### Breastfeeding outcomes

We examined five breastfeeding outcomes including initiation, establishment, sustained breastfeeding, exclusive breastfeeding duration and overall breastfeeding duration. Initiation was defined as receipt of any breast milk within the first 4 weeks after birth, compared with never breastfeeding or breastfeeding for less than 1 week. Establishment was defined as still breastfeeding at 2 months, and sustained breastfeeding as still breastfeeding at 6 months. Exclusive breastfeeding duration captured the period during which the infant received only breast milk, whereas overall duration captured the period during which the infant received any breast milk, including alongside other foods or liquids.

Outcomes other than initiation were assessed only among mothers who initiated breastfeeding. Breastfeeding information was obtained from maternal self-report questionnaires in ALSPAC, MoBa and BiB. In UKB, mothers were classified according to whether they had breastfed their offspring after maternal–offspring pairs were identified. Details of measurement, coding variables and derivation are provided in Supplementary Table S2 and Supplementary Material.

### Genotyping

Genotyping, imputation and genetic quality control were performed within each cohort using cohort-specific protocols. Analyses were restricted to participants of European ancestry. Relatedness, and sample- and SNP (single-nucleotide polymorphism)-level quality control were assessed before GWAS. Details of genotyping platforms, reference panels, and quality-control thresholds are provided in the Supplementary Material.

## Statistical analysis

### Genetic associations with breastfeeding outcomes

To obtain novel SNP–breastfeeding outcome associations for MR, we modelled cohort-specific maternal genotype GWAS using REGENIE,^20^ with logistic regression for binary outcomes and linear regression after inverse normal transformation (INT) for continuous outcomes, adjusting for maternal age, cohort-specific genetic principal components, and MoBa genotyping batch effects. Cohort-specific summary statistics were quality controlled, harmonized and meta-analyzed using METAL.^21^ Heterogeneity was assessed using Cochran’s Q and I² statistics. Corresponding offspring genotype GWAS were harmonized and meta-analyzed using the same procedures.

### Genetic associations with maternal and perinatal exposures

We investigated 17 exposures based on clinical interest and relevance for maternal and child health (Supplementary Table S3). These included sociodemographic exposures (e.g., educational attainment), cardiometabolic exposures (e.g., BMI, T2D, smoking, and blood pressures), psychiatric exposures (e.g., depression and insomnia), and perinatal exposure (e.g., gestational duration and birthweight). SNP–exposure association estimates were extracted from publicly available large-scale GWAS summary statistics limited on participants of European descent, to minimize population stratification bias (Supplementary Table S3). For binary exposures, genetic instruments are to be interpreted as indicating liability or propensity for the exposure.

### Genetically-predicted effects of maternal exposures on breastfeeding outcomes

We employed the inverse variance weighted (IVW) MR method^22^ to estimate (genetically predicted) effects of 17 exposures on 5 breastfeeding outcomes (Figure 1A), under the key assumptions that genetic instruments are associated with the exposure, independent of confounders, and influence breastfeeding only through the exposure (Supplementary Figure S1). SNPs were selected as genetic instruments from exposure GWAS at P<5×10−08 and clumped in PLINK 1.9^23^ to retain independent SNPs (pairwise linkage disequilibrium r^2^<0.01 using the 1000 Genomes European ancestry reference panel). SNP–exposure and SNP–breastfeeding associations were harmonized to the same effect allele.

### Mediation of the education–breastfeeding association

We used two-step MR mediation analysis (Figure 1B) to identify targetable mediators between educational attainment and breastfeeding outcomes. We assessed six candidate mediators previously identified as targetable exposures for optimizing pregnancy health:^24^ BMI, WHR, smoking, T2D, insomnia and depression. First, we estimated the effect of educational attainment on each mediator; second, we estimated the effect of each mediator on breastfeeding outcomes. The mediated effect was calculated as the product of these estimates, and the proportion mediated as the mediated effect divided by the total effect of educational attainment on breastfeeding. Standard errors for mediated effects were estimated using the delta method.^25^

### Evidence triangulation, robustness checks and bias assessments

We evaluated the robustness of the main results through complementary analyses that accounted for offspring genotype, compared MR estimates with confounder-adjusted multivariable regression models (MVA), and performing a range of MR sensitivity analyses (Figure 1C).

#### Adjusting for offspring genotype

To assess potential bias from offspring (and therefore paternal) genotype, we additionally adjusted the breastfeeding GWAS analyses for offspring genotype, through a weighted linear model (WLM)^26^ combining maternal and offspring GWAS estimates while accounting for overlapping samples. We then used these new genetic signals for breastfeeding in new, offspring-adjusted MR analyses. This was done for 3 of the breastfeeding traits and not for exclusive and sustained breastfeeding, for which offspring genotype data were unavailable in UKB and BiB.

#### Multivariable regression analysis

We used MVA in MoBa to compare observational associations with MR estimates, because MoBa had data on most exposures and all breastfeeding outcomes. Twelve of the 17 exposures were available for these analyses; WHR, high cholesterol, DBP and PTSD were unavailable in MoBa and were assessed only in MR analyses. Each breastfeeding outcome was regressed on each exposure, adjusting for covariates (Supplementary Table S4). Logistic regression was used for binary outcomes, and linear regression for INT continuous breastfeeding duration outcomes. Exposure definitions and measurement in MoBa are provided in the Supplementary Material.

#### MR sensitivity analyses

We conducted MR sensitivity analyses using methods that make different assumptions about horizontal pleiotropy, including MR Egger regression,^27^ MR PRESSO,^28^ the weighted median^29^ and weighted mode;^30^ tested heterogeneity and directional pleiotropy using Cochran’s Q and the MR-Egger intercept; and assessed instrument strength using F-statistics. We also compared combined-sex and female-only instruments where available (Supplementary Table S5) and evaluated potential sample overlap between exposure and outcome GWAS. Details are provided in the Supplementary Material.

Estimates were reported with 95% confidence intervals, and all tests were two-sided. MR estimates are presented as odds ratios (OR) for binary breastfeeding outcomes and coefficients (β) for continuous breastfeeding duration outcomes, interpreted per standard deviation (SD) difference after INT. Multiple testing was addressed within each analytical aim using the Benjamini–Hochberg false discovery rate (FDR). Analyses were conducted in R version 4.5.0 using TwoSampleMR v0.6.18, DONUTS, MR-PRESSO v1.0, ggplot2 v3.5.2 and metafor v4.8.0.

## Ethics and consent

All contributing studies had approval from relevant national or local ethics committees, and participants provided written informed consent. Cohort-specific ethics approvals are provided in the Supplementary Methods.

## Results

### Study populations and breastfeeding outcomes

The analyses included 72,653 mothers and 317,651 offspring from four cohorts in the UK and Norway (Supplementary Table S1). Cohort contributions varied by breastfeeding outcome as available breastfeeding measures differed. ALSPAC and MoBa contributed to all five outcomes, BiB to initiation, establishment and overall duration, and UKB to initiation only. Initiation rate was high across cohorts (Supplementary Figure S2), consistent with patterns in high-income countries. Continuation rates were higher in MoBa than in the UK-based cohorts, ALSPAC and BiB, which showed similar patterns. By 2 months postpartum, breastfeeding prevalence had declined to around 60% in ALSPAC and BiB and 80% in MoBa. At 6 months, sustained breastfeeding was reported by 28% of mothers in ALSPAC and 68% in MoBa.

### Genetic associations with breastfeeding outcomes

We generated SNP–breastfeeding outcome associations through cohort-specific GWAS and meta-analysis. Maternal-genotype meta-analyses showed limited between-cohort heterogeneity, with mean I² values of approximately 15–16% and around 5% of SNPs showing Cochran’s Q P<0.05 across outcomes (Supplementary Table S6). Several GWAS loci were identified for breastfeeding initiation, sustained breastfeeding and exclusive breastfeeding duration in maternal-genotype analyses, whereas offspring-genotype GWAS identified loci only for breastfeeding initiation (Supplementary Figures S3–S4).

### Genetically-predicted effects of maternal exposures on breastfeeding outcomes

We estimated effects of 17 exposures on five breastfeeding outcomes using MR. Eight exposures showed evidence of an effect on at least one breastfeeding outcome using the primary IVW MR method (Figure 2; Supplementary Figure S5). After FDR correction, educational attainment, BMI and WHR showed consistent evidence of effects across all breastfeeding outcomes; educational attainment, BMI, WHR, smoking and depression showed evidence for four outcomes.

**Figure 2.**
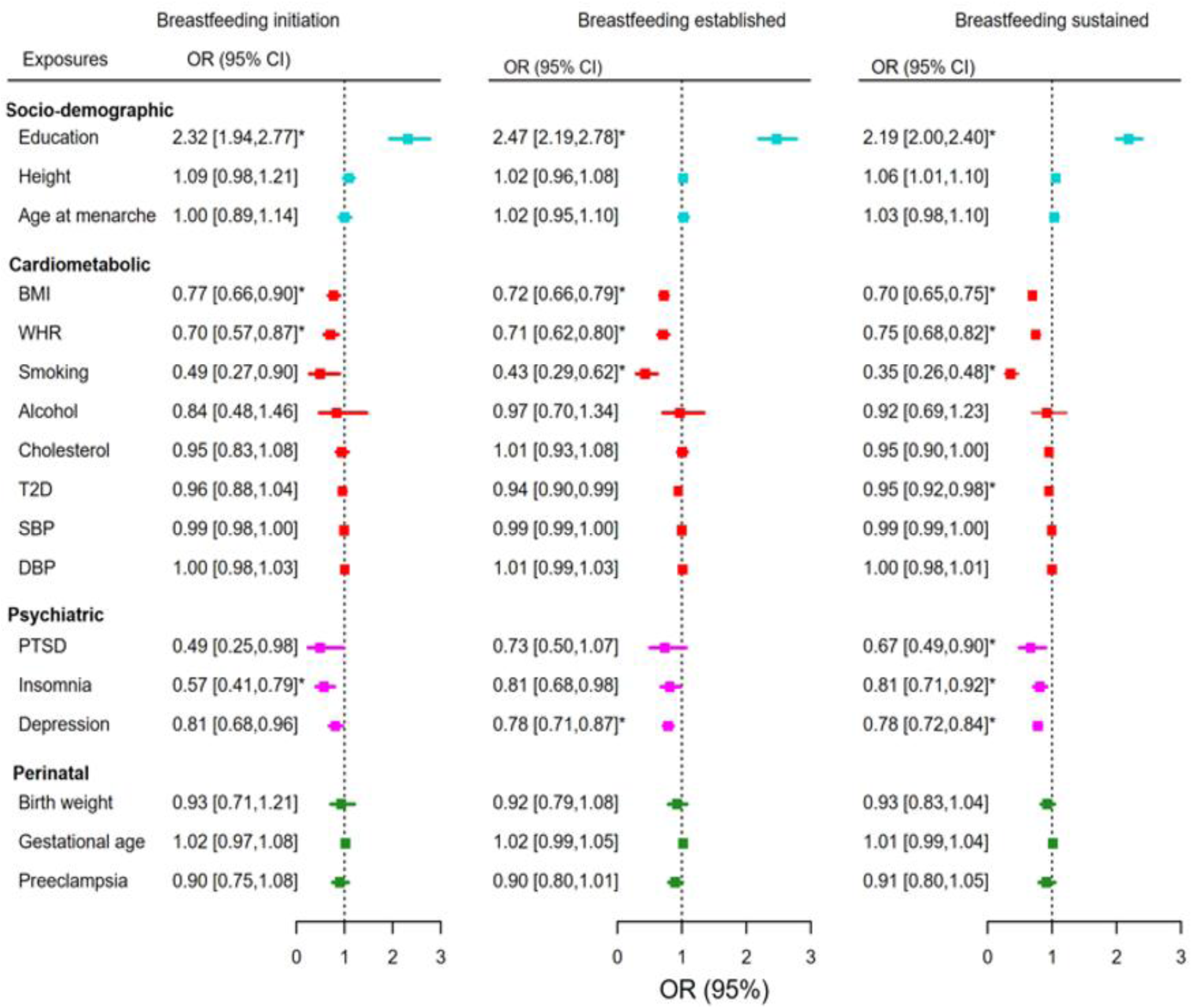
MR estimates for binary breastfeeding outcomes. Causal effects of 17 maternal and perinatal exposures on breastfeeding initiation, establishment and sustained breastfeeding were estimated using inverse variance weighted (IVW) MR method. Odds ratios (ORs) are shown with 95% confidence intervals; the vertical dotted line indicates the null effect (OR=1). Exposures are grouped by domain. Asterisks indicate associations significant after FDR correction.

Educational attainment showed positive effects, whereas cardiometabolic and psychiatric factors, including BMI, WHR, smoking, T2D, insomnia and depression, showed effects in the opposite direction. Estimates were broadly consistent across breastfeeding initiation and continuation. A 1SD (3.4-year) increase in educational attainment increased the odds of initiating (OR: 2.32, 95% CI: 1.94,2.77), establishing (OR: 2.47, 95% CI: 2.19,2.78) and sustaining breastfeeding (OR: 2.19, 95% CI: 2.00,2.40), and increased exclusive (β: 0.21SD, 95% CI: 0.17,0.24) and overall breastfeeding duration (β: 0.24SD, 95% CI: 0.22,0.27).

A 1SD increase in BMI lowered the odds of initiation (OR: 0.77, 95% CI: 0.66,0.90), establishment (OR: 0.72, 95% CI: 0.66,0.79) and sustained breastfeeding (OR: 0.70, 95% CI: 0.65,0.75), and reduced overall and exclusive breastfeeding duration by 0.12SD (β: -0.12, 95% CI: -0.16, -0.09) and 0.09SD (β: -0.09, 95% CI: -0.12, -0.06), respectively. WHR showed similar adverse associations, lowering breastfeeding odds by 25–30% and shortening duration. Smoking, insomnia and depression showed adverse effects across several breastfeeding outcomes, while PTSD showed evidence for breastfeeding initiation and overall duration. There was little evidence that genetically-predicted height, age at menarche, alcohol intake, cholesterol, SBP, DBP or perinatal exposures affected breastfeeding (Supplementary Table S7).

### Modifiable pathways linking maternal education to breastfeeding outcomes

We assessed whether BMI, WHR, smoking, T2D, insomnia and depression mediated the effect of educational attainment on breastfeeding outcomes. Results included the total effect, mediated effect and proportion mediated (PM), and are shown in Figure 3 for binary outcomes and Supplementary Figure S6 for continuous duration outcomes. BMI, WHR and smoking showed the strongest evidence of mediation. In contrast, T2D and depression showed more modest evidence, whereas insomnia showed no evidence of mediation. The proportion mediated ranged from 2% for exclusive breastfeeding duration, mediated by T2D alone (effect = 0.01, 95% CI: 0.00,0.01), to 29% for sustained breastfeeding, mediated by smoking alone (OR = 1.25, 95% CI: 1.17,1.34).

**Figure 3.**
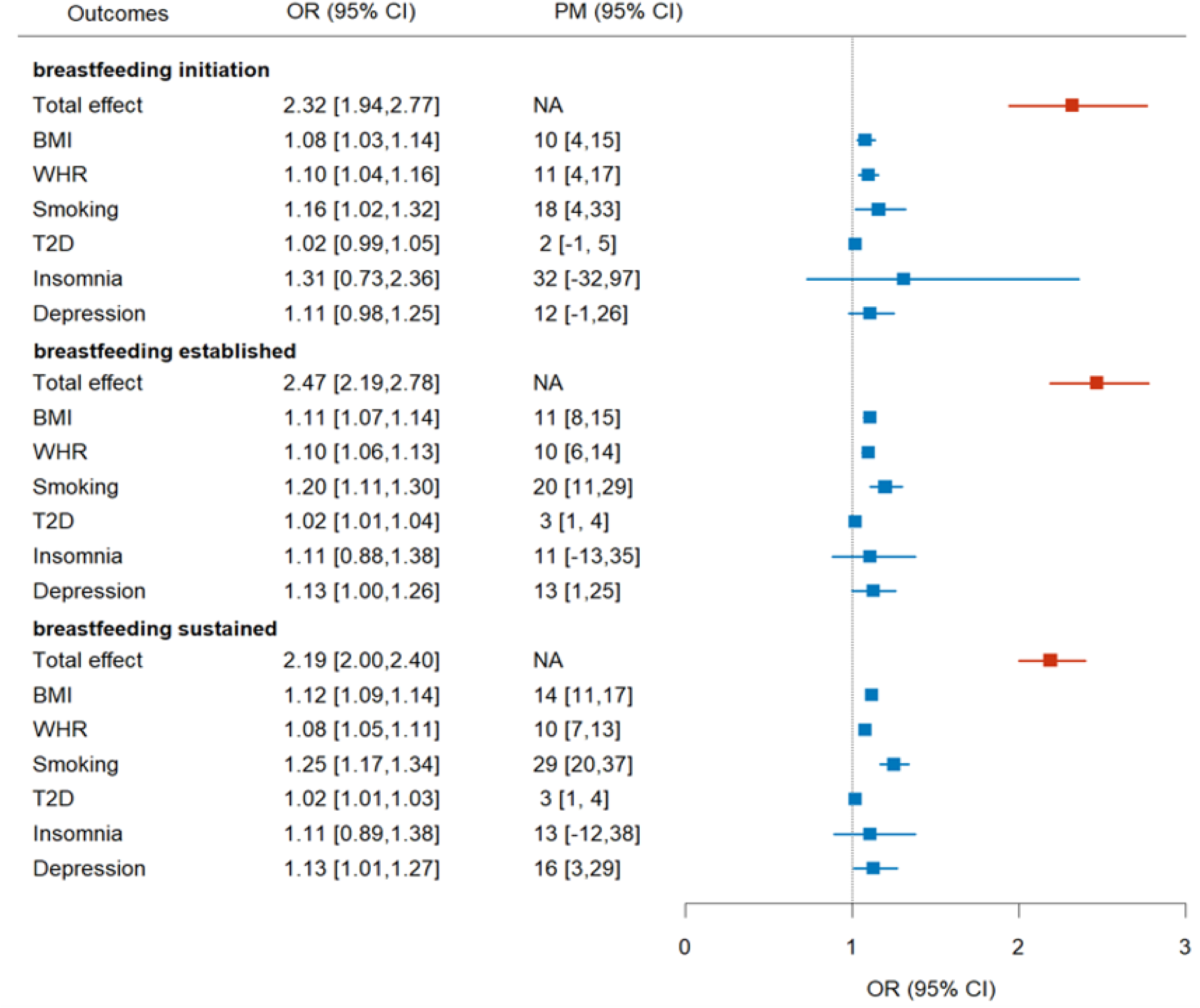
Mediation of the education–breastfeeding association. Two-step MR estimates of cardiometabolic and psychiatric mediation in the association between educational attainment and breastfeeding outcomes. Red indicates the total effect of education on breastfeeding outcomes, and light blue indicates the indirect effect through each candidate mediator. The figure also shows the proportion mediated (PM). NA indicates that PM was not estimated because there was no total effect of education on that breastfeeding outcome.

BMI mediated 19% of the education effect on overall duration (mediated effect = 0.04, 95% CI: 0.03,0.05), reducing to 10% for breastfeeding initiation (mediated OR = 1.08, 95% CI: 1.03,1.14). WHR showed a similar pattern, mediating 15% of the education effect on overall duration and 9% for initiation. Depression mediated up to 17% of the education effect on overall duration (mediated effect = 0.04, 95% CI: 0.00,0.07) and 13% for established breastfeeding (mediated OR = 1.13, 95% CI: 1.00,1.26).

### Triangulating results, robustness checks and bias assessment

#### Offspring-genotype adjustment and multivariable regression comparison

MR estimates accounting for offspring genotype and MVA estimates are presented in Figure 4. MR estimates adjusted for offspring genotype were less precise, with wider 95% confidence intervals, but were largely consistent in magnitude with unadjusted MR results. Some attenuation was observed for smoking and depression, particularly for breastfeeding initiation. Effects on established breastfeeding and overall duration were comparable between unadjusted and offspring genotype–adjusted MR.

**Figure 4.**
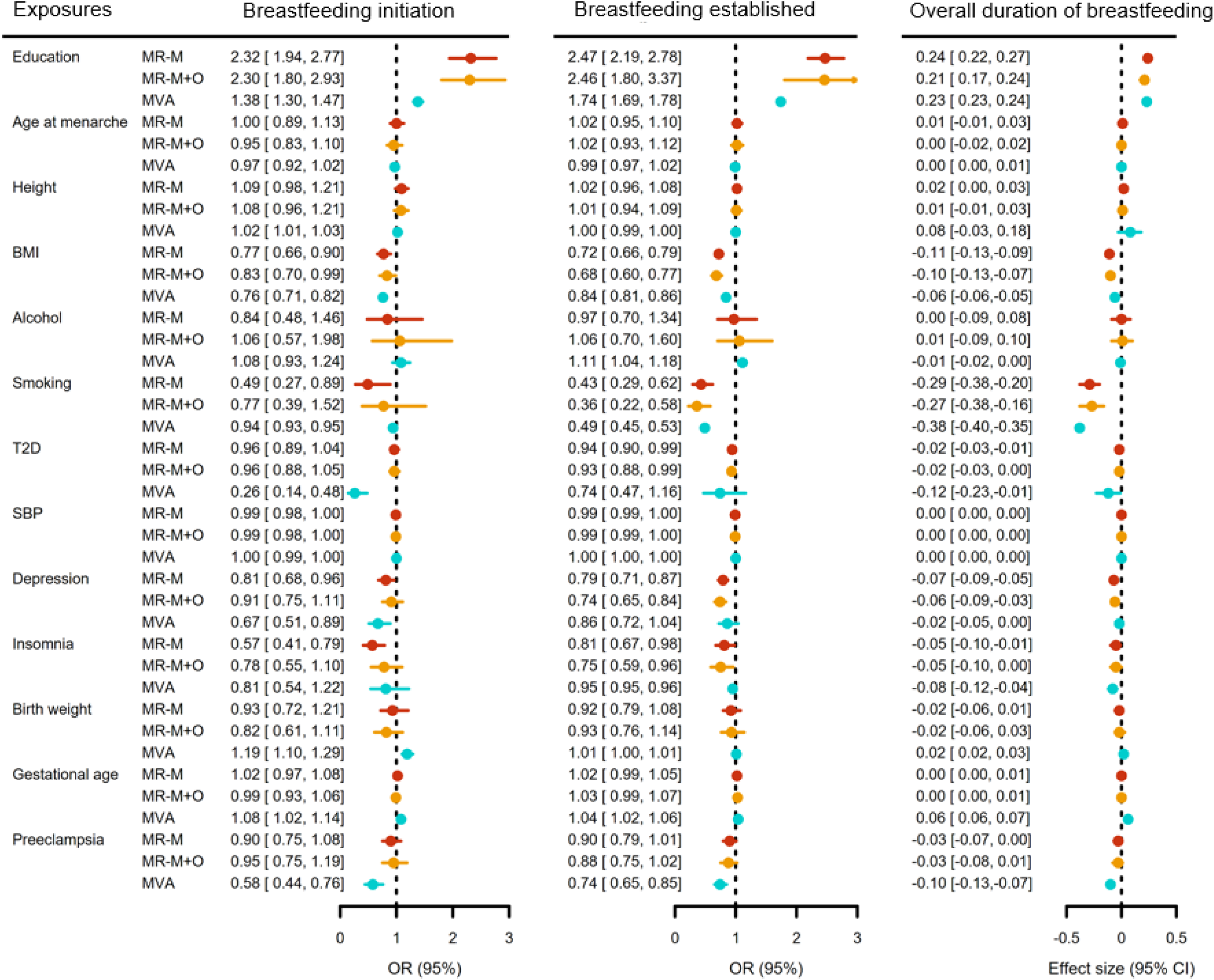
Offspring-genotype adjustment and multivariable regression comparison. Effect estimates are shown for primary MR using maternal genotype only (MR-M), MR adjusted for offspring genotype (MR-M+O), and multivariable regression (MVA) for three breastfeeding outcomes. For breastfeeding initiation and establishment, estimates are odds ratios (ORs) with 95% CIs; the vertical dotted line indicates the null effect (OR=1). For overall breastfeeding duration, estimates are coefficients (β) with 95% CIs; the vertical dotted line indicates the null effect (β=0).

MVA results generally aligned with MR, showing associations of educational attainment, BMI and smoking with breastfeeding outcomes. However, effect sizes diverged for some exposures. For example, depression was associated with 19% lower odds of breastfeeding initiation in MR, compared with a 33% reduction in MVA. Consistent with MR, MVA found no evidence for associations of age at menarche, alcohol intake or SBP with breastfeeding. Both approaches supported an association of T2D with shorter breastfeeding duration, but only MVA supported lower odds of breastfeeding initiation and establishment. In contrast to MR, MVA indicated positive associations of perinatal traits, including gestational age and preeclampsia, with breastfeeding outcomes. Comparisons were not possible for sustained breastfeeding and exclusive duration, which were available in the offspring GWAS for only two cohorts.

#### MR sensitivity analyses

Genetic instruments were strong across exposures, with F-statistics exceeding 10 (Supplementary Table S8). We observed instrument heterogeneity for sustained breastfeeding and either exclusive or overall breastfeeding duration (Cochran’s Q P < 0.05), but not for other key outcomes (Supplementary Table S9). There was no evidence of directional pleiotropy based on the MR-Egger intercept test (Supplementary Table S10). IVW results were generally consistent with alternative MR methods, with effects of the eight exposures supported by at least two of four MR methods (Supplementary Figures S7–S8; Supplementary Table S7). For mediation analyses, education-to-mediator estimates were largely consistent across MR methods, with no evidence of heterogeneity or directional pleiotropy (Supplementary Figure S9; Supplementary Table S11). We also compared combined-sex and female-only instruments, available for 10 of 17 exposures. Female-only instrument sets were smaller and estimates less precise, but results were directionally consistent with combined-sex analyses (Supplementary Figures S10–S11). Effect sizes were comparable for most exposures, including BMI and SBP, while educational attainment, WHR and smoking showed slight differences.

## Discussion

We provided robust insights to inform breastfeeding support strategies, by triangulating the evidence on maternal and perinatal determinants of breastfeeding success using four population-based large-scale cohorts and combining analytical approaches relying on different assumptions. We showed that lower educational attainment and higher BMI, smoking, T2D, insomnia and depression lead to suboptimal breastfeeding to varying degrees, and little evidence for a causal role of maternal height, age at menarche, blood pressure, cholesterol or perinatal exposures, including birth weight and gestational age. Causal mediation analyses indicated that maternal cardiometabolic and psychiatric exposures, particularly BMI, smoking and depression, partly explained the effect of maternal education on breastfeeding. Confidence in these results is strengthened by consistent findings across questionnaire-based analyses adjusted for multiple confounders (MVA) and genetic associations (MR) including a broad set of sensitivity analyses assessing potential assumptions violations.

Previous studies have consistently reported lower breastfeeding rates in less educated women in high-income countries,^6,31^ which our findings support. By leveraging genetic instruments, our results point to a causal relationship. Mediation analyses further indicated that modifiable factors (BMI, smoking and depression) partly explain the adverse effects of lower education on breastfeeding. This suggests that interventions targeting these exposures could help reduce breastfeeding disparities related to sociodemographic inequality. Our findings for BMI are consistent with observational evidence^7,15^ and a recent MR study limited to breastfeeding initiation.^15^ We extend this evidence across multiple breastfeeding outcomes, showing that both overall adiposity (BMI) and central adiposity (WHR) may affect breastfeeding beyond initiation. This could reflect metabolic interference with lactation, greater challenges establishing breastfeeding among women with higher BMI, or both. Similarly, genetic liability to T2D was associated with lower odds of establishing and sustaining breastfeeding and with shorter exclusive and overall breastfeeding duration, helping clarify inconsistent observational evidence reporting either no association^32^ or difficulties establishing exclusive breastfeeding.^9^ Smoking also showed adverse effects on breastfeeding outcomes, consistent with our MoBa regression results and previous observational studies.^8^ For psychiatric traits, genetically predicted depression and insomnia were associated with reduced breastfeeding duration, aligning with clinical evidence linking perinatal depression and sleep disturbance to breastfeeding difficulties.^4,10^ Although PTSD estimates were less precise, they were consistent with previous evidence that maternal trauma and PTSD may hinder sustained breastfeeding.^11^

Several mechanisms may explain these findings. Higher BMI may impair lactation through a diminished prolactin response,^33^ delayed lactogenesis and premature breastfeeding cessation.^34^ T2D- related insulin impairment may also disrupt mammary gland insulin signaling needed for secretory differentiation and milk synthesis.^35^ Smoking may act through nicotine transfer into breast milk, altered milk composition and disrupted infant feeding behaviour.^36^ Psychiatric conditions may affect breastfeeding through disruption of the oxytocin system, which supports milk ejection and maternal bonding.^37^ Depression has been linked to reduced oxytocin during lactation and feeding difficulties,^4^ while insomnia may worsen fatigue, and sleep disruption.^38^ These interpretations should be considered in light of gene–environment equivalence.^39^ For adiposity, smoking and depression, genetically proxied differences plausibly reflect biological or behavioral pathways affecting lactation and breastfeeding continuation. For education, interpretation is less direct; estimates likely reflect education-related social, behavioral and health pathways shaping breastfeeding success, rather than schooling alone.^40^ Some inconsistencies between MR and MVA were observed. For preeclampsia, birth weight and gestational age, MR provided little evidence of causal effects, whereas MVA and previous observational studies suggested associations with breastfeeding.^12,13^ This discrepancy may reflect limited MR power, as few SNPs were available for perinatal exposures, or residual confounding in observational analyses. If MR assumptions hold, these findings suggest that some observational associations for perinatal exposures may be partly confounded.

This study has several strengths. First, it is the largest to triangulate complementary genetic and observational evidence on maternal determinants of breastfeeding, combining data from 72,653 mothers across four cohorts. Second, we used large-scale GWAS to construct genetic instruments for a broad range of modifiable exposures and structural determinants, enabling a more holistic assessment than previous studies. Third, we incorporated maternal–offspring genotype adjustment, mediation analyses and complementary multivariable models, providing convergent evidence to strengthen causal inference. Finally, breastfeeding success was characterized using five distinct outcomes, extending beyond the conventional ever/never classification and enabling a more detailed assessment of breastfeeding patterns.

Limitations include that, although genetic instruments were strong, they were few for some exposures, reducing precision and limiting sensitivity analyses. Some breastfeeding outcomes were unavailable in all cohorts, reducing power and potentially contributing to minor inconsistencies across outcomes. We could not attempt bidirectional MR due to the lack of strong genetic instruments for breastfeeding outcomes. Second, some MR assumptions might not hold in all analyses. Horizontal pleiotropy remains possible, particularly for multifactorial exposures such as educational attainment, although consistent findings across sensitivity analyses suggest it is unlikely to meaningfully explain our results. Third, retrospective recall of breastfeeding in UKB may introduce noise through reporting bias.

However, primary analyses relied mainly on prospectively collected breastfeeding data from pregnancy and birth cohorts. Fourth, breastfeeding is highly complex, likely introducing more measurement noise than physiologically defined outcomes such as lactation insufficiency.

Nevertheless, we examined multiple dimensions of breastfeeding success, including establishment among women who initiated breastfeeding, a plausible proxy for lactation failure. Finally, analyses were restricted to women of European ancestry, limiting generalizability to populations with different genetic architectures and social determinants of breastfeeding. Future studies should replicate these analyses in more diverse populations using ancestry-specific instruments.

Finally, our findings provide evidence that maternal cardiometabolic and psychiatric exposures influence breastfeeding success and partly mediate educational differences in breastfeeding. They support monitoring and improving maternal health from the preconception period onwards, including cardiometabolic health, smoking cessation and mental health, as part of breastfeeding support strategies. They also suggest opportunities to identify women at higher risk of breastfeeding difficulties after initiation who may benefit from targeted support. Future research should test whether interventions addressing these pathways improve breastfeeding continuation and reduce inequalities, with potential long-term benefits for maternal and child health.

## Supporting information

Additional Supplementary Methods and Results

## Data Availability

Supporting data for breastfeeding outcomes are not publicly available due to data access restrictions and participant confidentiality. Researchers may apply for access through the respective cohort data access procedures subject to approval.
Instrumental variables for the exposures were obtained from publicly available GWAS summary statistics.

## Contributors

MWA, CB, ZK, and LZ contributed to the conceptualization of the study. MWA and LZ developed the analysis plan, performed the data analyses, and wrote the original draft of the manuscript. CB, MCM, NM, and RJH contributed to formal analyses of cohort-specific data and data acquisition. CB, MCM, NM, RJH, ZK, and LZ critically revised and edited the manuscript. All authors read and approved the manuscript for submission.

## Data sharing

Supporting data for breastfeeding outcomes are not publicly available due to data access restrictions and participant confidentiality. Researchers may apply for access through the respective cohort data access procedures: UK Biobank (https://www.ukbiobank.ac.uk/enable-your-research/apply-for-access), the Norwegian Mother, Father and Child Cohort Study (MoBa; https://www.fhi.no/en/studies/moba/for-researchers/), the Avon Longitudinal Study of Parents and Children (ALSPAC; https://www.bristol.ac.uk/alspac/researchers/access/), and Born in Bradford (BiB; https://borninbradford.nhs.uk/research/how-to-access-data/), subject to approval. Instrumental variables for the exposures were obtained from publicly available GWAS summary statistics (Supplementary Table S1).

## Acknowledgements

This research was conducted using the UKB resource under Application Number 102297. We are extremely grateful to all families who took part in ALSPAC, the midwives who helped recruit them, and the wider ALSPAC team, including data collection, administrative, technical and laboratory staff at the University of Bristol. ALSPAC data were obtained under Project ID B3346. We are grateful to all families participating in MoBa, which is supported by the Norwegian Institute of Public Health. MoBa data were obtained under Project ID p582. Born in Bradford is only possible because of the enthusiasm and commitment of the children and parents in BiB; we thank all participants, health professionals, schools and researchers who have made Born in Bradford happen. Details on study-specific acknowledgements are provided in Supplementary Material. We also thank Albert Navarro Gallinad and Laura Ghirardi for valuable feedback on the manuscript and helpful discussions.

## Code availability

Code used to perform the analyses and generate the figures in this study is made available at https://github.com/ht-diva/MR_exposure_breastfeeding-git.

## Supporting information

### Supplementary Material

Additional cohort details, genotyping and GWAS methods, exposure GWAS sources, MR implementation, mediation details, sensitivity analyses and additional results.

### Supplementary Figures

Supplementary Figures S1–S11

### Supplementary Tables

Supplementary Tables S1–S11

## References

1 Victora CG, Bahl R, Barros AJ, et al. Breastfeeding in the 21st century: epidemiology, mechanisms, and lifelong effect. The lancet 2016; 387: 475–90.

2 Miranda AR, Barral PE, Scotta AV, Cortez MV, Soria EA. An overview of reviews of breastfeeding barriers and facilitators: Analyzing global research trends and hotspots. Glob Epidemiol 2025; 9: 100192.

3 Scime NV, Shea AK, Faris P, Brennand EA. Impact of lifetime lactation on the risk and duration of frequent vasomotor symptoms: A longitudinal dose-response analysis. BJOG Int J Obstet Gynaecol 2023; 130: 89–98.

4 Nagel EM, Howland MA, Pando C, et al. Maternal Psychological Distress and Lactation and Breastfeeding Outcomes: a Narrative Review. Clin Ther 2022; 44: 215–27.

5 Vaz JS, Maia MFS, Neves PAR, Santos TM, Vidaletti LP, Victora C. Monitoring breastfeeding indicators in high-income countries: Levels, trends and challenges. Matern Child Nutr 2021; 17: e13137.

6 Neves PAR, Barros AJD, Gatica-Domínguez G, Vaz JS, Baker P, Lutter CK. Maternal education and equity in breastfeeding: trends and patterns in 81 low- and middle-income countries between 2000 and 2019. Int J Equity Health 2021; 20: 20.

7 Ballesta-Castillejos A, Gomez-Salgado J, Rodriguez-Almagro J, Ortiz-Esquinas I, Hernandez-Martinez A. Relationship between maternal body mass index with the onset of breastfeeding and its associated problems: an online survey. Int Breastfeed J 2020; 15: 55.

8 Liu J, Rosenberg KD, Sandoval AP. Breastfeeding Duration and Perinatal Cigarette Smoking in a Population-Based Cohort. Am J Public Health 2006; 96: 309–14.

9 Longmore DK, Barr ELM, Wilson AN, et al. Associations of gestational diabetes and type 2 diabetes during pregnancy with breastfeeding at hospital discharge and up to 6 months: the PANDORA study. Diabetologia 2020; 63: 2571–81.

10 Gordon LK, Mason KA, Mepham E, Sharkey KM. A mixed methods study of perinatal sleep and breastfeeding outcomes in women at risk for postpartum depression. Sleep Health 2021; 7: 353– 61.

11 Garthus-Niegel S, Horsch A, Ayers S, Junge-Hoffmeister J, Weidner K, Eberhard-Gran M. The influence of postpartum PTSD on breastfeeding: A longitudinal population-based study. Birth 2018; 45: 193–201.

12 Lutsiv O, Giglia L, Pullenayegum E, et al. A Population-Based Cohort Study of Breastfeeding According to Gestational Age at Term Delivery. J Pediatr 2013; 163: 1283–8.

13 Nejsum FM, Måstrup R, Torp-Pedersen C, Løkkegaard ECL, Wiingreen R, Hansen BM. Exclusive breastfeeding: Relation to gestational age, birth weight, and early neonatal ward admission. A nationwide cohort study of children born after 35 weeks of gestation. PLoS One 2023; **18**: e0285476.

14 Spry EA, Aarsman SR, Dashti SG, et al. Childhood sexual abuse and maternal social and financial resources, mental health, and parenting outcomes in pregnancy and early parenthood: a multicohort observational study. Lancet Obstet Gynaecol Women’s Health 2026; 2: e324–34.

15 Borges MC, Clayton GL, Freathy RM, et al. Integrating multiple lines of evidence to assess the effects of maternal BMI on pregnancy and perinatal outcomes. BMC Med 2024; 22: 32.

16 Ardissino M, Slob EAW, Carter P, et al. Sex-Specific Reproductive Factors Augment Cardiovascular Disease Risk in Women: A Mendelian Randomization Study. J Am Heart Assoc 2023; 12: e027933.

17 Davidsson OB, Rostgaard K, Kristjánsson RP, et al. Epidemiological and genetic evidence for shared mechanisms between migraine and pre-eclampsia: a nationwide cohort and genetic risk study in Denmark. Lancet Obstet Gynaecol Womens Health 2026; 2: e51–61.

18 Skrivankova VW, Richmond RC, Woolf BAR, et al. Strengthening the reporting of observational studies in epidemiology using mendelian randomisation (STROBE-MR): explanation and elaboration. BMJ 2021; 375: n2233.

19 Manichaikul A, Mychaleckyj JC, Rich SS, Daly K, Sale M, Chen W-M. Robust relationship inference in genome-wide association studies. Bioinformatics 2010; 26: 2867–73.

20 Mbatchou J, Barnard L, Backman J, et al. Computationally efficient whole-genome regression for quantitative and binary traits. Nat Genet 2021; 53: 1097–103.

21 METAL: fast and efficient meta-analysis of genomewide association scans. .

22 Burgess S, Butterworth A, Thompson SG. Mendelian Randomization Analysis With Multiple Genetic Variants Using Summarized Data. Genet Epidemiol 2013; 37: 658–65.

23 Chang CC, Chow CC, Tellier LC, Vattikuti S, Purcell SM, Lee JJ. Second-generation PLINK: rising to the challenge of larger and richer datasets. Gigascience 2015; 4: s13742–015.

24 Khan SS, Brewer LC, Canobbio MM, et al. Optimizing Prepregnancy Cardiovascular Health to Improve Outcomes in Pregnant and Postpartum Individuals and Offspring: A Scientific Statement From the American Heart Association. Circulation 2023; 147. DOI:10.1161/CIR.0000000000001124.

25 Carter AR, Sanderson E, Hammerton G, et al. Mendelian randomisation for mediation analysis: current methods and challenges for implementation. Eur J Epidemiol 2021; 36: 465–78.

26 Warrington NM, Hwang L-D, Nivard MG, Evans DM. Estimating direct and indirect genetic effects on offspring phenotypes using genome-wide summary results data. Nat Commun 2021; 12: 5420.

27 Bowden J, Davey Smith G, Burgess S. Mendelian randomization with invalid instruments: effect estimation and bias detection through Egger regression. Int J Epidemiol 2015; 44: 512–25.

28 Verbanck M, Chen C-Y, Neale B, Do R. Detection of widespread horizontal pleiotropy in causal relationships inferred from Mendelian randomization between complex traits and diseases. Nat Genet 2018; 50: 693–8.

29 Bowden J, Davey Smith G, Haycock PC, Burgess S. Consistent Estimation in Mendelian Randomization with Some Invalid Instruments Using a Weighted Median Estimator. Genet Epidemiol 2016; 40: 304–14.

30 Hartwig FP, Davey Smith G, Bowden J. Robust inference in summary data Mendelian randomization via the zero modal pleiotropy assumption. Int J Epidemiol 2017; 46: 1985–98.

31 Simpson DA, Carson C, Kurinczuk JJ, Quigley MA. Trends and inequalities in breastfeeding continuation from 1 to 6 weeks: findings from six population-based British cohorts, 1985–2010. Eur J Clin Nutr 2022; **76**: 671–9.

32 Laine MK, Kautiainen H, Gissler M, Pennanen P, Eriksson JG. Impact of gestational diabetes mellitus on the duration of breastfeeding in primiparous women: an observational cohort study. Int Breastfeed J 2021; 16: 19.

33 Grattan DR, Pi XJ, Andrews ZB, et al. Prolactin Receptors in the Brain during Pregnancy and Lactation: Implications for Behavior. Horm Behav 2001; 40: 115–24.

34 Preusting I, Brumley J, Odibo L, Spatz DL, Louis JM. Obesity as a Predictor of Delayed Lactogenesis II. J Hum Lact 2017; 33: 684–91.

35 Neville MC, Webb P, Ramanathan P, et al. The insulin receptor plays an important role in secretory differentiation in the mammary gland. Am J Physiol-Endocrinol Metab 2013; 305: E1103–14.

36 Napierala M, Mazela J, Merritt TA, Florek E. Tobacco smoking and breastfeeding: effect on the lactation process, breast milk composition and infant development. A critical review. Environ Res 2016; 151: 321–38.

37 Stuebe AM, Grewen K, Meltzer-Brody S. Association Between Maternal Mood and Oxytocin Response to Breastfeeding. J Womens Health 2013; 22: 352–61.

38 Mariman A, Hanoulle I, Pevernagie D, et al. Longitudinal assessment of sleep and fatigue according to baby feeding method in postpartum women: a prospective observational study. BMC Pregnancy Childbirth 2024; 24: 529.

39 Hemani G, Stender S, Wolters FJ, Hofman A, Davey Smith G. The rapid growth in Mendelian randomization studies. Eur J Epidemiol 2025; 40: 1165–71.

40 Howe LJ, Rasheed H, Jones PR, et al. Educational attainment, health outcomes and mortality: a within-sibship Mendelian randomization study. Int J Epidemiol 2023; 52: 1579–91.

