## Additional Supplementary Figures for "Maternal health factors driving breastfeeding success: an evidence triangulation framework"

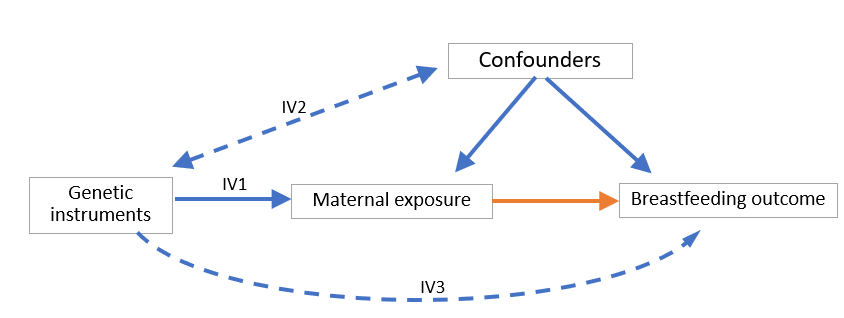

**Supplementary Figure S1**: Schematic presentation of the Mendelian Randomization (MR) design. Orange arrow represents the causal effect of interest. For MR estimate of causal effect to be valid, three main assumptions need to be met: IV1 the genetic variant is associated with the exposure; IV2: the genetic variant is not associated with outcome through a confounder; and IV3: the genetic variant does not directly affect the outcome.

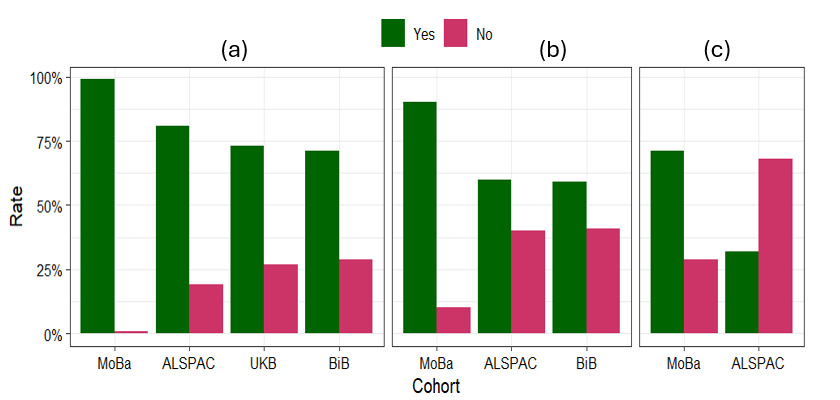

**Supplementary Figure S2:** Breastfeeding distributions across cohorts**.** Distribution of binary breastfeeding outcomes across cohorts. (a): breastfeeding initiation, (b): established breastfeeding at two months, (c): sustained breastfeeding at six months.

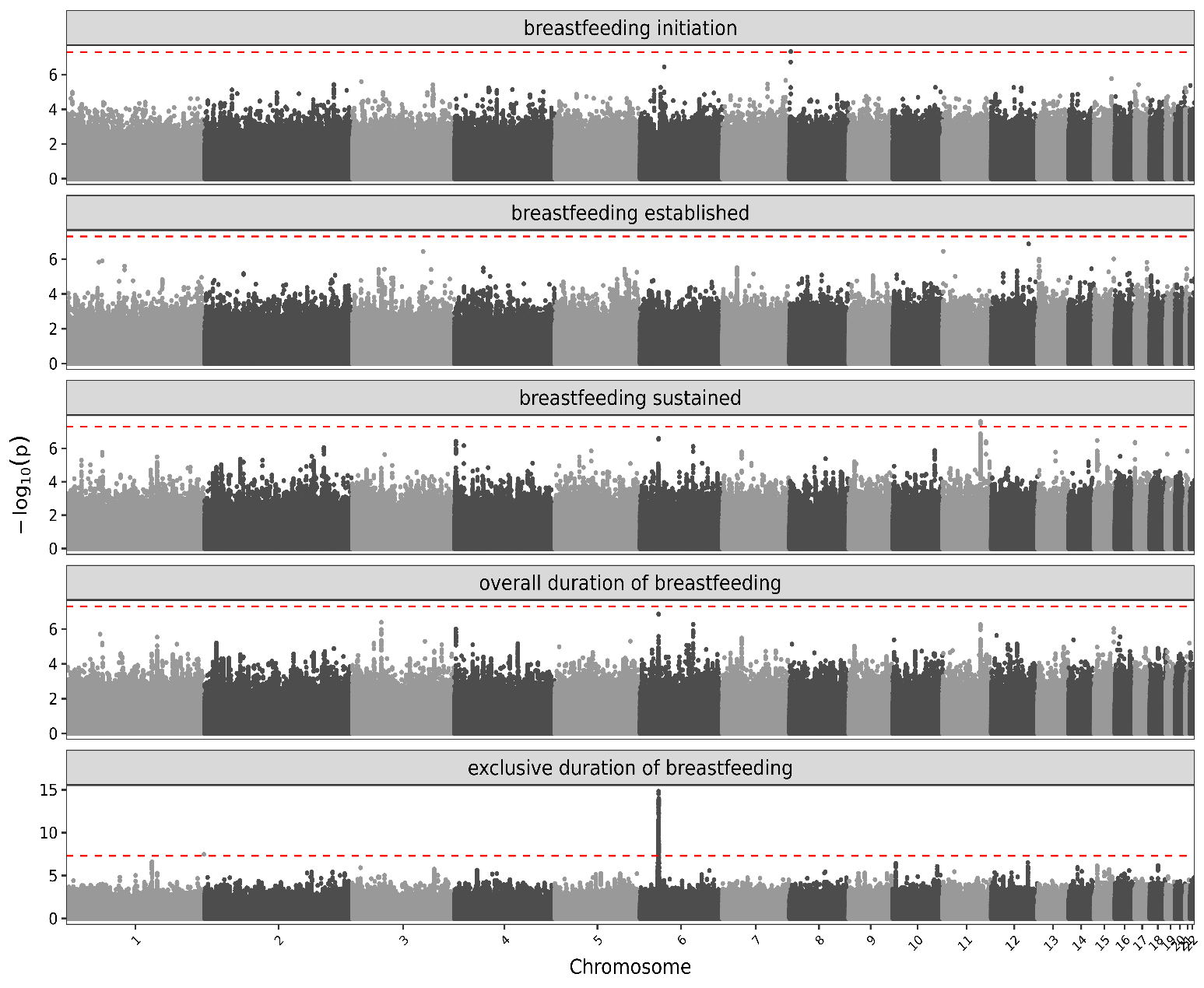

**Supplementary Figure S3**: Maternal-genotype GWAS results**.** Manhattan plots show genome-wide association results for five breastfeeding outcomes using maternal genotype data from 72,653 women across four European cohorts. Each panel represents one breastfeeding outcome. SNPs are plotted by autosomal genomic position, with the y-axis showing −log₁₀(P). Cohort-specific estimates were meta-analysed using fixed-effect inverse-variance weighting. The dashed line indicates genome-wide significance (P=5×10⁻⁸).

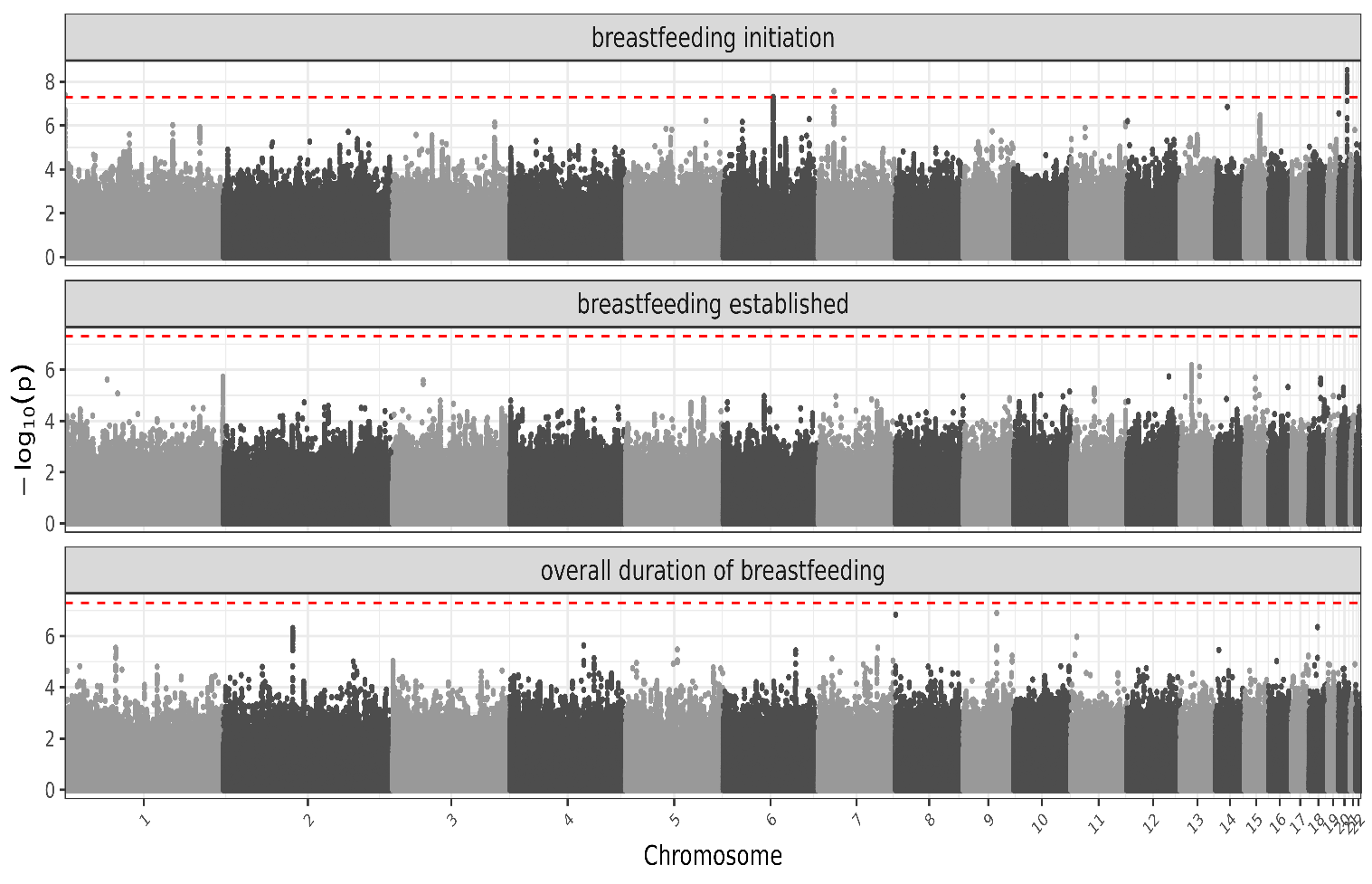
 **Supplementary Figure S4:** Offspring-genotype GWAS results**.** Manhattan plots show genome-wide association results for breastfeeding outcomes using offspring genotype data from 317,651 individuals across four European cohorts. Each panel represents one breastfeeding outcome. SNPs are plotted by autosomal genomic position, with the y-axis showing −log₁₀(P). Cohort-specific estimates were meta-analysed using fixed-effect inverse-variance weighting. The dashed line indicates genome-wide significance (P=5×10⁻⁸).

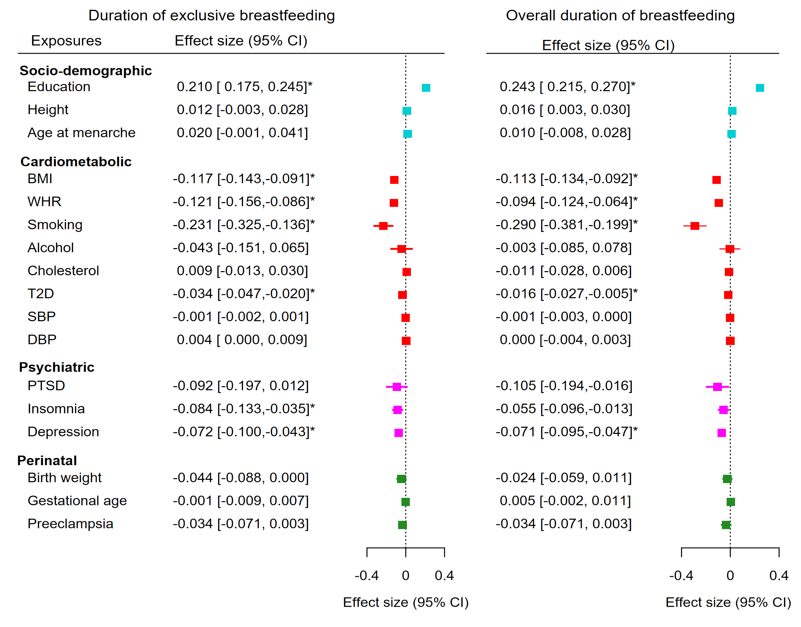

**Supplementary Figure S5:** MR estimates for continuous breastfeeding outcomes**.**
Causal effects of 17 maternal and perinatal exposures on exclusive and overall breastfeeding duration were estimated using inverse variance weighted Mendelian randomization. Estimates are shown with 95% confidence intervals; the vertical dotted line indicates the null effect (β=0). Exposures are grouped by domain. Asterisks indicate associations significant after FDR correction.

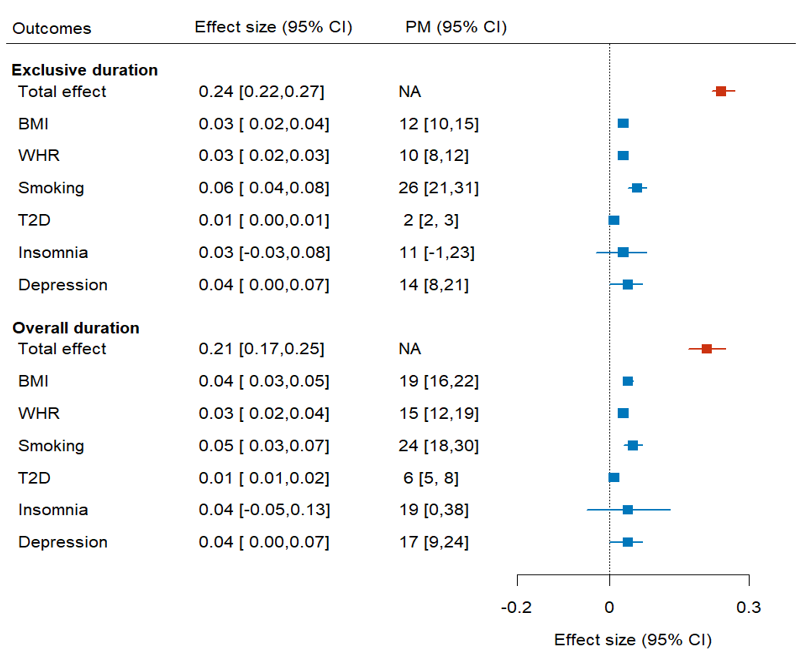

**Supplementary Figure S6: Mediation of the education–breastfeeding association for continuous breastfeeding outcomes.**Two-step MR estimates of cardiometabolic and psychiatric mediation in the association between educational attainment and exclusive and overall breastfeeding duration. Red indicates the total effect of education on breastfeeding duration, and light blue indicates the indirect effect through each candidate mediator. The figure also shows the proportion mediated (PM). NA indicates that PM was not estimated because there was no total effect of education on that breastfeeding outcome.

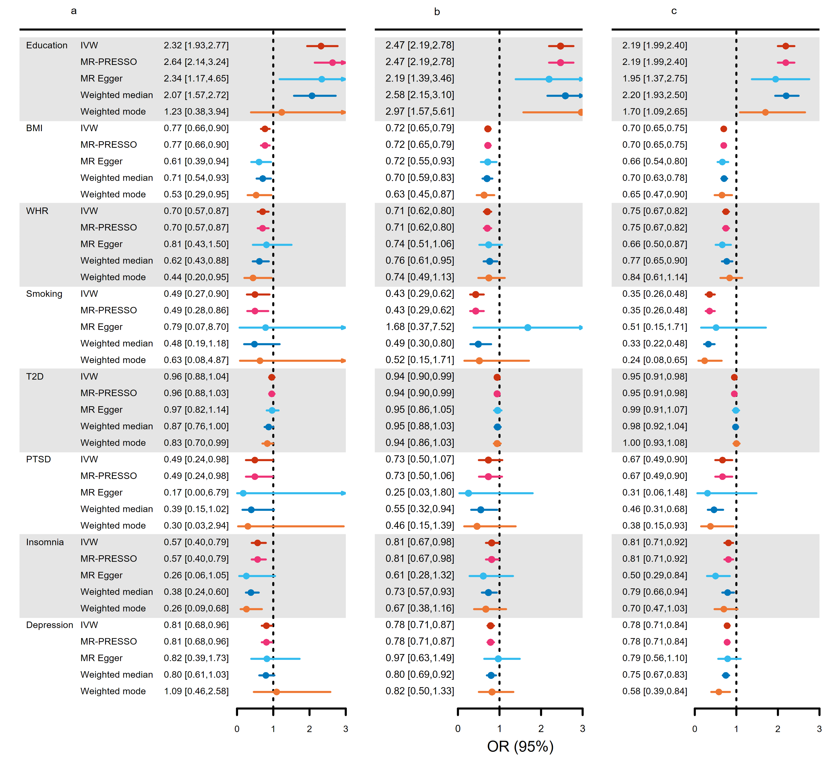

**Supplementary Figure S7:** MR sensitivity analyses for binary breastfeeding outcomes.
MR sensitivity estimates for the eight exposures that showed evidence of association with at least one breastfeeding outcome in the primary IVW analysis. Effects on breastfeeding initiation, establishment and sustained breastfeeding were estimated using five MR methods: IVW, MR-PRESSO, MR-Egger, weighted median and weighted mode. Odds ratios (ORs) are shown with 95% confidence intervals; the vertical dotted line indicates the null effect (OR=1).

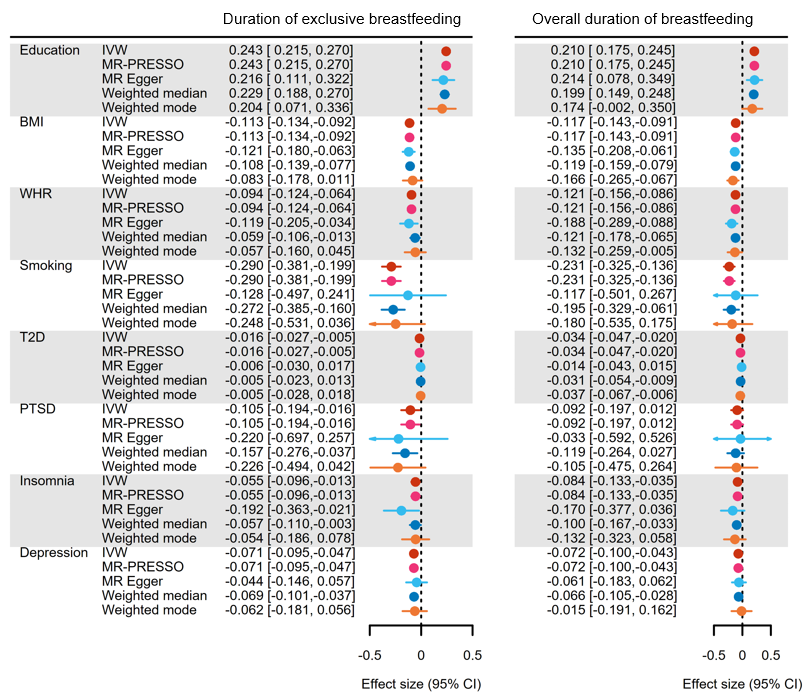

**Supplementary Figure S8:** MR sensitivity analyses for continuous breastfeeding outcomes. MR sensitivity estimates for the eight exposures that showed evidence of association with at least one breastfeeding outcome in the primary IVW analysis. Effects on exclusive and overall breastfeeding duration were estimated using five MR methods: IVW, MR-PRESSO, MR-Egger, weighted median and weighted mode. Estimates are shown with 95% confidence intervals; the vertical dotted line indicates the null effect (β=0).

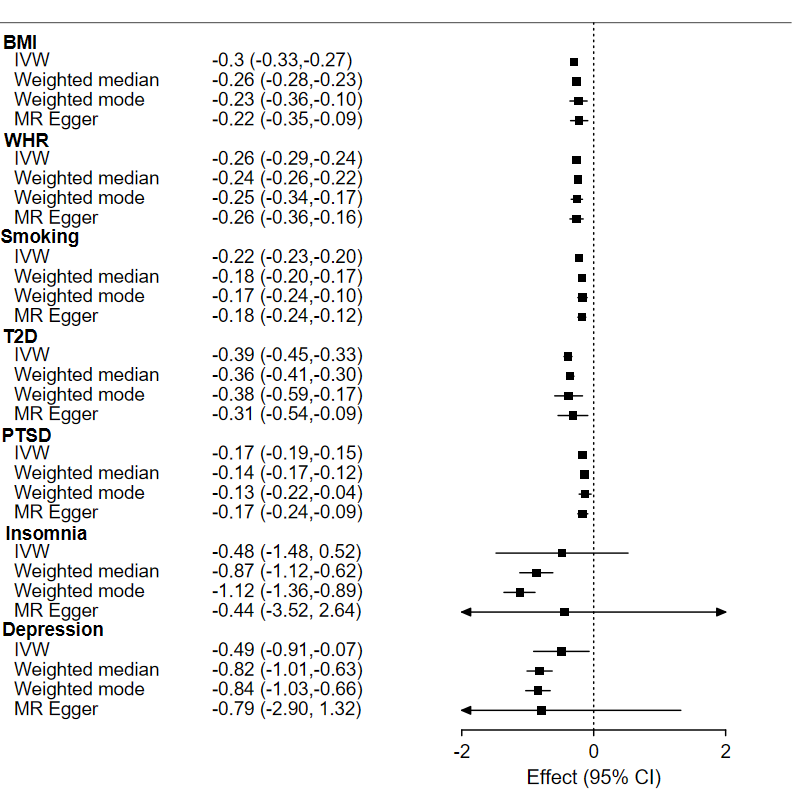

**Supplementary Figure S9:** Education-to-mediator MR estimates. Two-sample MR estimates for the association of genetically predicted educational attainment with cardiometabolic and psychiatric mediators. For each mediator, estimates are shown using IVW, weighted median, weighted mode and MR-Egger methods. IVW estimates were used to calculate the mediated, or indirect, effect of education on breastfeeding outcomes through each mediator.

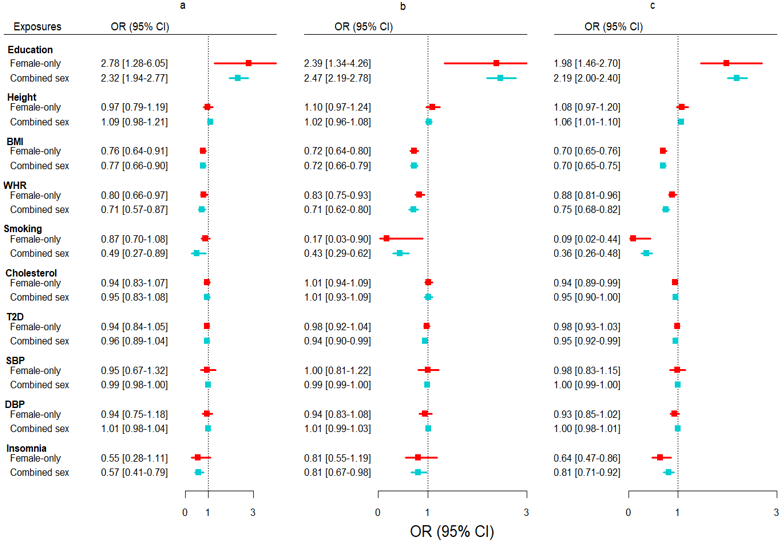

**Supplementary Figure S10:** Combined-sex versus female-only instruments for binary breastfeeding outcomes. MR estimates for 10 exposures using genetic instruments derived from combined-sex versus female-only GWAS. Effects are shown for breastfeeding initiation (a), establishment (b) and sustained breastfeeding (c). Odds ratios (ORs) are presented with 95% confidence intervals; the vertical dotted line indicates the null effect (OR=1).

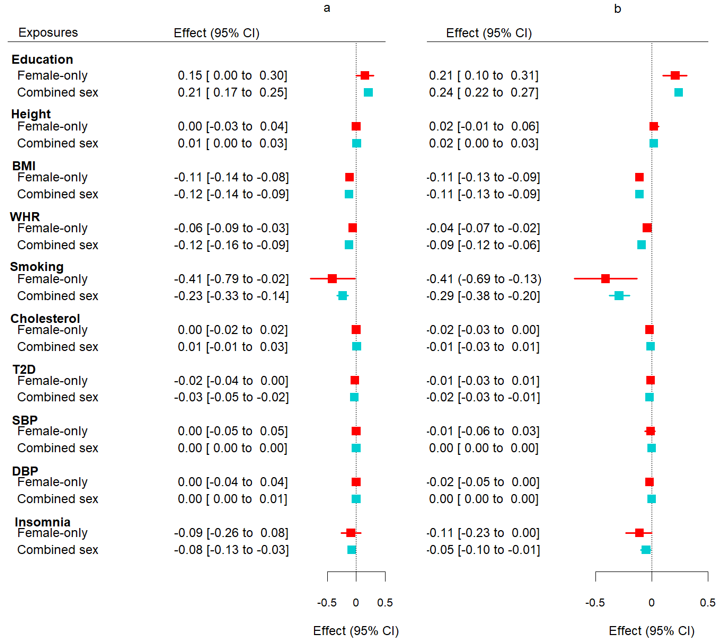
 **Supplementary Figure S11:** Combined-sex versus female-only instruments for continuous breastfeeding outcomes. MR estimates for 10 exposures using genetic instruments derived from combined-sex versus female-only GWAS. Effects are shown for exclusive and overall breastfeeding duration. Estimates are presented with 95% confidence intervals; the vertical dotted line indicates the null effect (β=0).
