## Additional Supplementary Methods and Results for "Maternal health factors driving breastfeeding success: an evidence triangulation framework"

**Supplementary Materials**

**Supplementary Methods**

**Study populations and cohort-specific description**

**UK Biobank (UKB):** The UKB is a prospective cohort study that enrolled participants at ages 40–49 years. A full study protocol is described elsewhere.^1^ In brief, all people in the UK National Health Service registry within the age range and living within a 25 mile radius from one of 22 study centres were invited to participate between 2006-2010. In total 501, 258 adults were recruited into UKB. Participants who had a valid email address (N=339,229) were invited to fill in a detailed online questionnaire assessing their early life after January 2015, and 158,835 participants fully completed it by October 2017.^2^

*Ethical approval in UKB*

Ethical approval for UKB was obtained and our study was performed under UKB application number 102297.

*Genotyping, imputation and quality control (QC) in UKB*

Genotyping, pre-imputation QC, and imputation procedures were described in detail elsewhere,^3^ and are briefly summarized here. UKB men and women were genotyped on two arrays. The first ~50,000 samples were genotyped on the UK BiLEVE array and the remaining ~450000 samples were genotyped on the UK Biobank Axiom array. Genotype data were imputed against two references panels: Haplotype Reference Consortium (HRC) panel and UK10K + 1000 Genomes panel. We used the imputed data released by UKB in March 2018, and we applied post-imputation QC, i.e. genetic sex same as reported sex, XX or XY in sex chromosome and no outliers in heterozygosity and missing rates).

*Breastfeeding outcome data in UKB*

At the assessment centres, participants completed a touchscreen questionnaire providing information on early life factors. Breastfeeding data were collected with the question “Were you breastfed when you were a baby” (yes/no/do not know/prefer not to answer). Additional variables used for phenotype data cleaning include maternal smoking around birth (yes/no), participant year of birth, ethnicity (re-categorised into Black, Asian, White, mixed, other), birthweight (kg), if adopted (yes/no), participant age at enrolment (years). Participants were excluded if they answered, “do not know”, “prefer not to answer", had missing data or provided inconsistent response between the baseline and the two follow-ups for the question if they were breastfed as infant. Additionally, participants were excluded if they were part of a multiple births, adopted. A total of 247000 sample with qualified breastfeeding information passed the QC procedure.

*Maternal-Offspring pair in UKB*

The UKB has no direct maternal breastfeeding information as it was not a birth cohort study, the presence of thousands of family members provides an opportunity to build maternal cohort by inferring the parent-offspring pair using genotype data. We used 501, 258 genotyped UK Biobank samples of British or Irish ancestry, together with their pairwise kinship estimates, to identify a subset of samples with mothers. To identify these pairs, we used pairwise kinship and identity by descent estimates using the KING^4^ and provided as part of the UKB study. We also added the condition of age difference greater than 15 years between mother-offspring pair. We used the age and sex of the individuals to distinguish mothers and offspring. We found a total of 4176 mother-offspring pairs within the full dataset, consistent with the previous parent-offspring findings.^3–5^

**Avon Longitudinal Study of Parents and Children (ALSPAC)**

ALSPAC is a geographically based prospective cohort study investigating the health and development of children. The study protocol was described elsewhere.^6,7^ Briefly, pregnant women resident in Avon, UK with expected dates of delivery 1^st^ April 1991 to 31^st^ December 1992 were invited to take part in the study. The initial number of pregnancies enrolled is 14,541 (for these at least one questionnaire has been returned or a ‘Children in Focus’ clinic data had been attended by 19/07/99).^6,7^ Of these initial pregnancies, there was a total of 14,676 foetuses, resulting in 14,062 live births and 13,988 children who were alive at 1 year of age.^6,7^ This study relied on 13,867 pregnancies from a total of 13,761 women, and most pregnancies were recruited at the first antenatal clinic visit in the first trimester of pregnancy.^6^ Questionnaires were sent at regular intervals during pregnancy. The study website contains details of all the data that is available through a fully searchable data dictionary and variable search tool (<http://www.bristol.ac.uk/alspac/researchers/our-data>).

*Ethical approval in ALSPAC*

Ethical approval for the study was obtained from the ALSPAC Ethics and Law Committee and the Local Research Ethics Committees. Consent for biological samples has been collected in accordance with the Human Tissue Act (2004). Informed consent for the use of data collected via questionnaires and clinics was obtained from participants following the recommendation of the ALSPAC Ethics and Law Committee at the time. Our study was performed under the ALSPAC data access approval application number B3346.

*Genotyping, imputation and QC in ALSPAC*

Genotyping, pre-imputation quality control, and imputation procedures were described in detail elsewhere,^8^ and are briefly summarized here. ALSPAC mothers were genotyped using Illumina human660K quad single nucleotide polymorphism (SNP) chip, and ALSPAC children were genotyped using Illumina HumanHap550 quad genome-wide SNP genotyping platform. Genotype data for both ALSPAC mothers and children were imputed against HRC v1.1 reference panel, after a similar QC procedure (minor allele frequency (MAF) ≥1%, a call rate ≥95%, in Hardy-Weinberg equilibrium (HWE), correct sex assignment, no evidence of cryptic relatedness, and of European descent).

*Breastfeeding outcome data in ALSPAC*

Breastfeeding data in ALSPAC were obtained from maternal self-reported questionnaires at approximately 1, 6, and 15 months postpartum. Responses were used to create five breastfeeding outcomes. Breastfeeding initiation: a categorical variable with breastfeeding initiated within the first month vs not initiated, which was defined using the questionnaire “Any breast feeding” (yes/no/missing) within the first month after birth. Next, duration of any breastfeeding was determined from 3 questionnaires: (1) latest report of breastfeeding at 4 weeks (1^st^ day/1^st^ week/2^nd^ week/3^rd^ week/4^th^ week/missing), (2) duration of breastfeeding reported at 6-month (never/less than a month/1-3 months/3-6 months/>6 months/missing), (3) age stopped breastfeeding reported at 15 months (never/still/missing). This indicated maximum number of months breast milk provided along with formula or other liquid/solid food. We used the binary classifications for breastfeeding established if breastfeeding for >=2months vs < 2months, and breastfeeding sustained if breastfeeding for >=6months vs <6months. The ALSPAC post-delivery questionnaires provided data on exclusive breastfeeding duration. Mothers were asked at what age the child started formula, follow-on milk, Soya milk, Goat milk, Cow and use of other bottle milk. The responses included age (in months), missing and not started yet. breastfeeding exclusive was then defined as being breastfed and not given any other drink of solid food in the time one is exclusively breastfed within 6 months after birth. Only live live-born singleton offspring were considered for the analyses. Further, participants were excluded for gestational duration less than 32 weeks and birth weight < 2.5 kg. Finally, N=6826 women of European descent with qualified genotype and breastfeeding data were eligible for inclusion in our MR analyses.

**The Norwegian Mother, Father and Child Cohort Study (MoBa)**

MoBa is a population-based pregnancy cohort study conducted by the Norwegian Institute of Public Health. Participants were recruited from all over Norway from 1999-2008.^9^ The cohort now includes 114,500 children, 106,889 mothers and 75,200 father.^9^ The current study is based on version 12 of the quality-assured data files released for research on “Prenatal environmental exposures and pregnancy outcomes-Mendelian randomization analysis”. The establishment of MoBa and initial data collection was based on a license from the Norwegian Data Protection Agency and approval from The Regional Committees for Medical and Health Research Ethics. The MoBa cohort is now based on regulations related to the Norwegian Health Registry Act. The Medical Birth Registry (MBRN) is a national health registry containing information about all births in Norway. MoBa has been linked to the Medical Birth Register of Norway (MBRN, established in 1967), using unique personal identification numbers.^9^ Blood samples were obtained from both parents during pregnancy and from mothers and children (umbilical cord) at birth.^10^

*Ethical approval in MoBa*

The current study was approved by The Regional Committees for Medical and Health Research Ethics. Access to MoBa data were obtained under Project ID p582.

*Genotyping, imputation and QC in MoBa*

Genotyping, pre-imputation quality control, and imputation procedures were described in detail elsewhere^11^ and we briefly summarized here. There were five projects that contributed to MoBa genetics 1.0,^12^ and we had to use an earlier version consisting of two projects – HARVEST and ROTTERDAM1 given their complete QC procedure. In HARVEST, MoBa mothers and children were genotyped using either Illumina HumanCoreExome12v1.1 or Illumina HumanCoreExome24v1.0. In ROTTERDAM1, MoBa mothers and children were genotyped using Illumina GSAMDv1.0. Genotype data from all batches were imputed against HRC v1.1 reference panel, after a similar QC procedure (MAF ≥5%, a call rate ≥99.2%, in HWE, correct sex assignment and no evidence of cryptic relatedness). Women of European descent with qualified genotype data and live-born singleton offspring were eligible for inclusion in our analyses (N=60,192).

*Breastfeeding outcome data in MoBa*

The MoBa self-reported questionnaires at 6 months post-delivery provided data on breastfeeding outcomes, where mothers were retrospectively reported what they had given their child to drink or eat for the first week and each month of the baby’s life. The questionnaire at 6 months includes questions: What did you give your child to drink during the first week of life, with response alternatives, breast milk, water, sugar water, formula, other (specify), don’t know/don’t remember. In addition, a question on: “What has your child been given to drink during the first 6 months of his/her life? The participants were to indicate for each month the child had been given the relevant drink (breast milk, standard Collett formula, Collett formula with Omega 3, Standard NAN formula, Nan HA1 formula, other milk (specify), water, squash/juice. breastfeeding initiation was defined as binary whether mothers provided any breastmilk during the first 4 weeks of life or not. For mothers who initiated breastfeeding, we defined the binary classifications breastfeeding established if breastfeeding >=2months vs < 2months, and breastfeeding sustained if breastfeeding for >=6months vs <6months. Overall duration of any breastfeeding was the maximum number of months for a mother who initiated breastfeeding reported that they exclusively or partially gave their child breastmilk up until the sixth month of life. To calculate exclusive breastfeeding outcome, additional self-reported information on solid food was used. Mothers were asked to report the age of the child when they started to provide him/her 16 types of solid food for the first time. Exclusive breastfeeding was then defined as being breastfed and not given any other drink of solid food in the time one is exclusively breast fed, as recommended by the 6-month WHO guidelines.^13^

*Maternal and perinatal exposure data in MoBa*

Maternal age at delivery was recorded in MBRN, and five women were grouped as “less than 17 years”. Therefore, we recoded them as “NA”. Age at menarche (in years) was self-reported at15 weeks of gestation. We excluded values below 7 and above 20 years as unlikely answers. Weight, height, the highest educational attainment were self-reported at 15 weeks of gestation. We excluded implausibly extreme values (weight less than 30 kg or more than 200 kg, height shorter than 135 cm). Various alternatives were given for maternal education (9- year secondary school, 1-2 year high school, technical school, 3-year high school general studies, junior college, technical college, 4-year university degree, more than 4 years (Master’s degree, medical doctor, PhD) and other education.

Maternal BMI was calculated from maternal weight and height as weight in kilograms divided by height in metres squared (kg/m²). Maternal smoking in pregnancy was self-reported at both 15 and 30 weeks of gestation. For each time point, we derived a binary smoker versus nonsmoker measure of smoking status by combining “daily” and “sometimes” smoking, as described previously.^14^ Smokers in pregnancy were participants who smoked at either 15 or 30 weeks of gestation, while nonsmokers were those who did not smoke at both time points. Alcohol intake during 0-12, 13-24 and 25-30 weeks of gestation were self-reported at 30 weeks of gestation, with seven levels from “never” to “roughly 6-7 times a week”, and we determined a binary exposed versus nonexposed measure of alcohol consumption during 0-30 weeks of gestation. Exposed group were participants who consumed alcohol at least once during the 30 weeks of gestation, while non-exposed group were those who never consumed at all time points. Systolic blood pressure and diastolic blood pressure levels were measured at 15 weeks of gestation. Diagnosis of Type 2 diabetes was self-reported at 15 weeks of gestation, recorded as a binary variable with type 2 diabetes cases (“yes”) versus control (“no”).

For the mental factors such as insomnia, MoBa participants were asked if they experienced sleeping problems during weeks 0-4, 5-8, 9-12, after 13 and 15 weeks of gestation of pregnancy (response “yes/no/missing”). For our analysis, we defined a binary variable of experiencing insomnia if a participant answered “yes” at least in one of the times versus never experienced insomnia if a participant answered “no” for all the times. Maternal depression was defined based on self-reported depression and recorded as answering “Yes” to having depression at 15 or 30 weeks, anxiety at 15 weeks or other psychological problems at 30 weeks, as previously done in other studies to define depressive disorder during pregnancy.^15^ Gage at delivery (weeks), birth weight (grams), and maternal preeclampsia status (any vs no preeclampsia) were obtained from MBRN.

**Born in Bradford (BiB) study**

The Born in Bradford (BiB) study is a population-based prospective birth cohort. In total, 12,453 women who had 13,776 pregnancies were recruited at ~24-28 weeks gestation at a routine oral glucose tolerance test (OGTT). At the time of recruitment, this was offered to all women booked for delivery at Bradford Royal Infirmary (BRI) (except for those with pre-existing diabetes (N = 70 - 0.5% of BiB pregnancies).^16^ All women recruited had an expected delivery between March 2007 and December 2010. Full details of the study methodology were reported previously.^17^ In brief, Bradford is a city in the North of England with high levels of socioeconomic deprivation. BiB has high proportions of White European and South Asian families, all residing in Bradford, UK, which makes it a unique cohort demographic to study. Parents (usually the mother) who were recruited into the study provided full informed consent, as well as detailed interview questionnaire data, measurements, and biological samples. They also consented to the linkage of their and their child’s data to routine (primary and secondary care) health and education data.

*Ethical approval in BiB*

Ethical approval for the study was granted by the Bradford National Health Service Research Ethics Committee (ref 06/Q1202/48).

*Breastfeeding outcome data in BiB*

Data on breastfeeding were extracted from the 6- and 12-month postnatal questionnaires included information on the initiation and duration of breastfeeding and the age at which formula milk, solid foods and drinks were introduced. Where data collected at both the 6- and 12-month visits were available, data collected at the 6-month visit were used in preference to those collected at the 12-month visit to reduce the likelihood of recall bias.^18^ Breastfeeding initiation was defined as any breastfeeding immediately after birth. This was recorded as a binary (yes/no) variable. Overall breastfeeding duration was defined as any breastfeeding duration providing any breast milk, regardless of supplementation with formula/foods. Breastfeeding established was defined as still breastfeeding at 2 months versus stopped before 2 months.

**Estimation of SNP–breastfeeding outcome associations**

To obtain SNP–breastfeeding outcome association estimates for Mendelian randomization, we conducted cohort-specific genome-wide association analyses (GWAS) for each breastfeeding outcome using maternal genotype data. GWAS were performed using REGENIE,^19^ which uses a two-step procedure: first fitting a whole-genome regression model to account for relatedness and population structure, and then testing individual variants for association with each outcome. Genetic variants were analysed as imputed allele dosages under an additive genetic model. Binary outcomes, including breastfeeding initiation, breastfeeding establishment, and sustained breastfeeding, were analysed using logistic regression. Continuous outcomes, including exclusive and overall breastfeeding duration, were analysed using linear regression after inverse normal transformation (INT). All models were adjusted for maternal age, cohort-specific genetic principal components (PC), and genotyping batch effects in MoBa.

Cohort-specific GWAS summary statistics were centrally quality controlled and harmonized before meta-analysis. Variants were aligned to a common genome build and effect allele, and variants with missing association statistics, inconsistent allele information, or poor imputation quality were excluded. Cohort-specific estimates were combined using fixed-effect meta-analysis as implemented in META. Between-cohort heterogeneity was assessed using Cochran’s Q and I² statistics. We also conducted corresponding cohort-specific GWAS using offspring genotype data for each breastfeeding outcome. These analyses used the same outcome definitions, genetic model, and cohort-specific quality control procedures as the maternal genotype GWAS, with adjustment for offspring sex, cohort-specific genetic PC and genotyping batch effects in MoBa. Offspring genotype GWAS results were harmonized and meta-analysed using the same procedures as described for the maternal genotype analyses.

**Exposure GWAS sources and genetic instruments**

We included 17 exposures based on clinical interest and relevance for maternal and child health (Supplementary Table S3). We extracted summary statistics from publicly available large-scale GWAS on each exposure including educational attainment,^20^ height,^21^ age at menarche,^22^ BMI,^23^ WHR,^23^ smoking,^24^ alcohol,^25^ total cholesterol,^26^ T2D,^27^ SBP,^28^ DBP,^28^ PTSD,^29^ insomnia,^30^ depression,^31^ preeclampsia,^32^ birth weight^33^ and gestational age.^34^ For binary exposures, genetic instruments are to be interpreted as indicating liability or propensity for the trait. For the perinatal exposures (gestational age and birth weight), genetic instruments were derived from maternal GWAS. Although offspring GWAS were available, they were not used, as they were not suitable for aim 1, which focused on estimating the causal effects of maternal exposures. To reduce potential bias due to population stratification, we restricted GWAS studies with participants of European descent.

**Triangulating results, robustness checks and bias assessment**

We evaluated the robustness of main results through complementary analyses that accounted for offspring genotype, compared MR estimates with confounder-adjusted multivariable regression models, and performing a range of MR sensitivity analyses.

*Adjusting for offspring genotype using a weighted linear model*

To adjust the maternal GWAS effects for effects of the offspring genotype, we applied a weighted linear model (WLM).^35^ This is because breastfeeding is a two-body phenotype directly involving both mother and offspring and effects of maternal variants might also be inherited by the offspring. The WLM estimates the adjusted maternal genetic effect as a linear combination of the unadjusted maternal and offspring genetic effects, conditional on overlapping samples as implemented in the DONUTS R package.^36^ Using these offspring-adjusted maternal GWAS estimates, we repeated the MR analysis for each exposure and breastfeeding outcome. These offspring-adjusted MR analyses were conducted for three breastfeeding outcomes (initiation, established and overall duration), as data on exclusive breastfeeding and sustained breastfeeding at 6 months were not available in UKB and BiB.

*Multivariable regression analysis*

Multivariable regression analysis (MVA) was used to estimate observational associations, which are more likely to be affected by biases such as confounding, between maternal exposures before/during pregnancy and breastfeeding outcomes in MoBa for comparison with our two-sample MR estimates. MoBa was the only cohort with available data on most of the 17 exposures included in the two-sample MR analyses. We included 12 maternal exposures (education, BMI, height, age at menarche, smoking, alcohol intake, SBP, T2D, insomnia, depression, gestational age and birth weight) measured during or before pregnancy. Four maternal exposures (WHR, high cholesterol, DBP and PTSD) were available only for MR analyses, as MoBa lacked measurements for MVA. Details on the definition, selection, and measurement of each exposure in the MoBa study are provided in Supplementary Method. We regressed each breastfeeding outcome on each maternal exposure adjusting for a set of covariates (Supplementary Table S4). We used logistic regression for the binary outcomes (breastfeeding initiation, established and sustained), and linear regression for the two continuous outcomes (exclusive and overall duration of breastfeeding) after normalizing the distributions using INT and cleaning as described for the GWAS analyses.

To ensure comparability between observational and Mendelian randomization estimates, the continuous exposures in the conventional regression analyses were transformed to match the scale used in the corresponding GWAS. Specifically, when GWAS summary statistics were based on standardized (z-transformed) traits, the exposure was likewise standardized in the observational analyses. This allowed effect estimates from both approaches to be interpreted per 1 standard deviation increase in the exposure, ensuring that comparisons were made on the same scale. Individuals with missing exposure, outcome or covariate data required for a given analysis were excluded from that analysis.

*MR sensitivity analyses*

We conducted a series of sensitivity analyses that relax the assumptions made about the nature of horizontal pleiotropy including MR Egger regression^37^, MR PRESSO^38^, the weighted median^39^ and weighted mode^40^. We tested between-SNP heterogeneity and directional pleiotropy in effect estimates using Cochran’s Q-statistic and the MR-Egger intercept test^37^, respectively. The strength of the selected instruments was assessed using F-statistics and proportion of variance explained by the instruments. Instrument selection for the main MR estimates were based on available large-scale GWAS conducted on all sex.

We evaluated weather these estimates substantially differed when genetic instruments were derived from female-only cohorts whenever sex-specific GWASs are available. Female-specific GWAS summary statistics were available for 10 of the 17 exposures analyzed (Supplementary Table S5), including educational attainment,^41^ height,^42^ BMI,^23^ WHR,^23^ smoking,^43^ total cholesterol,^26^ T2D,^27^ SBP,^44^ DBP^44^ and insomnia.^30^ Genetic instruments for combined-sex MR analyses were larger, included more instrumental variables, than those for female-only analyses across all exposures.

Finally, sample overlap between exposure and outcome GWAS was assessed using publicly available cohort descriptions (Supplementary Table S3). We found that UKB contributed to several exposure GWAS and to the breastfeeding initiation GWAS, so we excluded it from the initiation MR analyses when overlap occurred. ALSPAC and MoBa overlapped with some perinatal exposure GWAS (e.g. gestational age, birth weight). Although exact overlap could not be quantified, strong instrument strength suggests any overlap-induced bias is unlikely to meaningfully affect MR estimates.

**Results**

**Additional Mendelian randomisation results for maternal and perinatal exposures**

The estimated effects of smoking indicated markedly lower breastfeeding -reducing the odds of initiation by 50% (OR = 0.50, 95% CI: 0.27,0.90), establishing breastfeeding by 57% (OR = 0.43, 95% CI: 0.30,0.62), and sustaining breastfeeding by 63% (OR = 0.36, 95% CI: 0.26,0.48). Smoking was also estimated to reduce exclusive duration by 0.23SD (β: -0.23,95% CI: -0.32,-0.14) and overall duration by 0.29SD (β=-0.29, 95% CI: -0.38,-0.20). T2D effect estimates were similar across breastfeeding outcomes; however, the initiation estimate was less precisely estimated, with a wider confidence interval than the other outcomes.

Psychiatric traits showed a strong impact on breastfeeding. PTSD was linked to 51% lower odds of initiation (OR: 0.49, 95% CI: 0.25,0.98) and 0.1SD shorter overall duration of breastfeeding (β: -0.105, 95% CI: -0.194, -0.016). Insomnia was linked to 19% lower odds of both established (OR: 0.81, 95% CI: 0.68,0.98) and sustained (OR:0.81, 95% CI: 0.71,0.92), and 0.08SD and 0.05SD shorter exclusive (β: -0.08, 95% CI: -0.13, -0.03) overall (β: -0.05, 95% CI: -0.1, -0.01) breastfeeding duration. Depression was linked to 22% lower odds of both established (OR: 0.78, 95% CI: 0.71,0.87) and sustained (OR:0.78, 95% CI: 0.72,0.84), and 0.07SD shorter exclusive (β: -0.07, 95% CI: -0.1, –0.04) and overall (β: -0.07, 95% CI: -0.09, -0.047) duration of breastfeeding.

**Acknowledgment**

*UK Biobank:* The authors are grateful to UK Biobank participants and investigators for access to data to undertake this study (project number 102297).

*MoBa*: The Norwegian Mother, Father and Child Cohort Study is supported by the Norwegian Ministry of Health and Care Services and the Ministry of Education and Research. We are grateful to all the participating families in Norway who take part in this on-going cohort study. We thank the Norwegian Institute of Public Health (NIPH) for generating high-quality genomic data. This research is part of the HARVEST collaboration, supported by the Research Council of Norway (#229624). We also thank the NORMENT Centre for providing genotype data, funded by the Research Council of Norway (#223273), South East Norway Health Authorities and Stiftelsen Kristian Gerhard Jebsen. We further thank the Center for Diabetes Research, the University of Bergen for providing genotype data and performing quality control and imputation of the data funded by the ERC AdG project SELECTionPREDISPOSED, Stiftelsen Kristian Gerhard Jebsen, Trond Mohn Foundation, the Research Council of Norway, the Novo Nordisk Foundation, the University of Bergen, and the Western Norway Health Authorities”. This research has been conducted using MoBa data using Project ID p582

*ALSPAC:* We are extremely grateful to all the families who took part in this study, the midwives for their help in recruiting them, and the whole ALSPAC team, which includes interviewers, computer and laboratory technicians, clerical workers, research scientists, volunteers, managers, receptionists and nurses. Please note that the study website contains details of all the data that is available through a fully searchable data dictionary and variable search tool" and reference the following webpage: http://www.bristol.ac.uk/alspac/researchers/our-data/

*BiB*: Born in Bradford is only possible because of the enthusiasm and commitment of the children and parents in BiB. We are grateful to all the participants, health professionals and researchers who have made Born in Bradford happen.

**Funding**

*MoBa study funding:* MoBa is supported by the Norwegian Ministry of Health and Care services and the Ministry of Education and Research.

*ALSPAC study funding:* The UK Medical Research Council and Wellcome (Grant ref: 217065/Z/19/Z) and the University of Bristol provide core support for ALSPAC. A comprehensive list of grants funding is available on the ALSPAC website (http://www.bristol.ac.uk/alspac/external/documents/grant-acknowledgements.pdf); GWAS data was generated by Sample Logistics and Genotyping Facilities at Wellcome Sanger Institute and LabCorp (Laboratory Corporation of America) using support from 23andMe.

*BiB study funding:* Born in Bradford is supported by a Wellcome programme grant (WT223601/Z/21/Z: Age of Wonder), a Wellcome  infrastructure grant (WT101597MA), a joint Medical Research Council (MRC) and UK Economic and Social Science Research Council (ESRC) programme grant (MR/N024391/1) and a British Heart Foundation Clinical Study Grant (CS/16/4/32482). Funding for DNA extraction and genotyping was from two MRC programme grants (MC_UU_00011/6 and MC_UU_12013/5).

**References**

1	Collins R. What makes UK Biobank special? *The Lancet* 2012; **379**: 1173–4.

2	TouchscreenQuestionsMainFinal.pdf. https://biobank.ndph.ox.ac.uk/ukb/ukb/docs/TouchscreenQuestionsMainFinal.pdf (accessed Jan 4, 2025).

3	Bycroft C, Freeman C, Petkova D, *et al.* The UK Biobank resource with deep phenotyping and genomic data. *Nature* 2018; **562**: 203–9.

4	Manichaikul A, Mychaleckyj JC, Rich SS, Daly K, Sale M, Chen W-M. Robust relationship inference in genome-wide association studies. *Bioinformatics* 2010; **26**: 2867–73.

5	Hofmeister RJ, Rubinacci S, Ribeiro DM, Buil A, Kutalik Z, Delaneau O. Parent-of-Origin inference for biobanks. *Nat Commun* 2022; **13**: 6668.

6	Fraser A, Macdonald-Wallis C, Tilling K, *et al.* Cohort profile: the Avon Longitudinal Study of Parents and Children: ALSPAC mothers cohort. *Int J Epidemiol* 2013; **42**: 97–110.

7	Boyd A, Golding J, Macleod J, *et al.* Cohort profile: the ‘children of the 90s’—the index offspring of the Avon Longitudinal Study of Parents and Children. *Int J Epidemiol* 2013; **42**: 111–27.

8	Richmond RC, Timpson NJ, Felix JF, *et al.* Using genetic variation to explore the causal effect of maternal pregnancy adiposity on future offspring adiposity: a Mendelian randomisation study. *PLoS Med* 2017; **14**: e1002221.

9	Magnus P, Birke C, Vejrup K, *et al.* Cohort profile update: the Norwegian mother and child cohort study (MoBa). *Int J Epidemiol* 2016; **45**: 382–8.

10	Paltiel L, Anita H, Skjerden T, *et al.* The biobank of the Norwegian Mother and Child Cohort Study–present status. *Nor Epidemiol* 2014; **24**. https://www.ntnu.no/ojs/index.php/norepid/article/view/1755/1752 (accessed Jan 6, 2025).

11	Magnus MC, Miliku K, Bauer A, *et al.* Vitamin D and risk of pregnancy related hypertensive disorders: mendelian randomisation study. *bmj* 2018; **361**. https://www.bmj.com/content/361/bmj.k2167.abstract (accessed Jan 6, 2025).

12	Projects that have contributed to MoBa Genetics. GitHub. https://github.com/folkehelseinstituttet/mobagen/wiki/Projects-that-have-contributed-to-MoBa-Genetics (accessed Jan 6, 2025).

13	Breastfeeding. https://www.who.int/health-topics/breastfeeding (accessed Jan 6, 2025).

14	Yang Q, Borges MC, Sanderson E, *et al.* Associations between insomnia and pregnancy and perinatal outcomes: Evidence from mendelian randomization and multivariable regression analyses. *PLoS Med* 2022; **19**: e1004090.

15	Olstad EW, Nordeng HME, Sandve GK, Lyle R, Gervin K. Effects of prenatal exposure to (es)citalopram and maternal depression during pregnancy on DNA methylation and child neurodevelopment. *Transl Psychiatry* 2023; **13**: 1–11.

16	Taylor K, McBride N, Goulding NJ, *et al.* Metabolomics datasets in the Born in Bradford cohort. *Wellcome Open Res* 2021; **5**: 264.

17	Wright J, Small N, Raynor P, *et al.* Cohort Profile: The Born in Bradford multi-ethnic family cohort study. *Int J Epidemiol* 2013; **42**: 978–91.

18	Santorelli G, Fairley L, Petherick ES, Cabieses B, Sahota P. Ethnic differences in infant feeding practices and their relationship with BMI at 3 years of age – results from the Born in Bradford birth cohort study. *Br J Nutr* 2014; **111**: 1891–7.

19	Mbatchou J, Barnard L, Backman J, *et al.* Computationally efficient whole-genome regression for quantitative and binary traits. *Nat Genet* 2021; **53**: 1097–103.

20	Okbay A, Wu Y, Wang N, *et al.* Polygenic prediction of educational attainment within and between families from genome-wide association analyses in 3 million individuals. *Nat Genet* 2022; **54**: 437–49.

21	Yengo L, Sidorenko J, Kemper KE, *et al.* Meta-analysis of genome-wide association studies for height and body mass index in ∼700000 individuals of European ancestry. *Hum Mol Genet* 2018; **27**: 3641–9.

22	Day FR, Thompson DJ, Helgason H, *et al.* Genomic analyses identify hundreds of variants associated with age at menarche and support a role for puberty timing in cancer risk. *Nat Genet* 2017; **49**: 834–41.

23	Pulit SL, Stoneman C, Morris AP, *et al.* Meta-analysis of genome-wide association studies for body fat distribution in 694 649 individuals of European ancestry. *Hum Mol Genet* 2019; **28**: 166–74.

24	Wootton RE, Richmond RC, Stuijfzand BG, *et al.* Evidence for causal effects of lifetime smoking on risk for depression and schizophrenia: a Mendelian randomisation study. *Psychol Med* 2020; **50**: 2435–43.

25	Saunders GRB, Wang X, Chen F, *et al.* Genetic diversity fuels gene discovery for tobacco and alcohol use. *Nature* 2022; **612**: 720–4.

26	Graham SE, Clarke SL, Wu K-HH, *et al.* The power of genetic diversity in genome-wide association studies of lipids. *Nature* 2021; **600**: 675–9.

27	Mahajan A, Taliun D, Thurner M, *et al.* Fine-mapping type 2 diabetes loci to single-variant resolution using high-density imputation and islet-specific epigenome maps. *Nat Genet* 2018; **50**: 1505–13.

28	Keaton JM, Kamali Z, Xie T, *et al.* Genome-wide analysis in over 1 million individuals of European ancestry yields improved polygenic risk scores for blood pressure traits. *Nat Genet* 2024; **56**: 778–91.

29	Nievergelt CM, Maihofer AX, Atkinson EG, *et al.* Genome-wide association analyses identify 95 risk loci and provide insights into the neurobiology of post-traumatic stress disorder. *Nat Genet* 2024; **56**: 792–808.

30	Watanabe K, Jansen PR, Savage JE, *et al.* Genome-wide meta-analysis of insomnia prioritizes genes associated with metabolic and psychiatric pathways. *Nat Genet* 2022; **54**: 1125–32.

31	Adams MJ, Streit F, Meng X, *et al.* Trans-ancestry genome-wide study of depression identifies 697 associations implicating cell types and pharmacotherapies. *Cell* 2025; **188**: 640-652.e9.

32	Tyrmi JS, Kaartokallio T, Lokki AI, *et al.* Genetic Risk Factors Associated With Preeclampsia and Hypertensive Disorders of Pregnancy. *JAMA Cardiol* 2023; **8**: 674–83.

33	Warrington NM, Beaumont RN, Horikoshi M, *et al.* Maternal and fetal genetic effects on birth weight and their relevance to cardio-metabolic risk factors. *Nat Genet* 2019; **51**: 804–14.

34	Solé-Navais P, Flatley C, Steinthorsdottir V, *et al.* Genetic effects on the timing of parturition and links to fetal birth weight. *Nat Genet* 2023; **55**: 559–67.

35	Warrington NM, Hwang L-D, Nivard MG, Evans DM. Estimating direct and indirect genetic effects on offspring phenotypes using genome-wide summary results data. *Nat Commun* 2021; **12**: 5420.

36	Wu Y, Zhong X, Lin Y, *et al.* Estimating genetic nurture with summary statistics of multigenerational genome-wide association studies. *Proc Natl Acad Sci* 2021; **118**: e2023184118.

37	Bowden J, Davey Smith G, Burgess S. Mendelian randomization with invalid instruments: effect estimation and bias detection through Egger regression. *Int J Epidemiol* 2015; **44**: 512–25.

38	Verbanck M, Chen C-Y, Neale B, Do R. Detection of widespread horizontal pleiotropy in causal relationships inferred from Mendelian randomization between complex traits and diseases. *Nat Genet* 2018; **50**: 693–8.

39	Bowden J, Davey Smith G, Haycock PC, Burgess S. Consistent Estimation in Mendelian Randomization with Some Invalid Instruments Using a Weighted Median Estimator. *Genet Epidemiol* 2016; **40**: 304–14.

40	Hartwig FP, Davey Smith G, Bowden J. Robust inference in summary data Mendelian randomization via the zero modal pleiotropy assumption. *Int J Epidemiol* 2017; **46**: 1985–98.

41	Okbay A, Beauchamp JP, Fontana MA, *et al.* Genome-wide association study identifies 74 loci associated with educational attainment. *Nature* 2016; **533**: 539–42.

42	Randall JC, Winkler TW, Kutalik Z, *et al.* Sex-stratified Genome-wide Association Studies Including 270,000 Individuals Show Sexual Dimorphism in Genetic Loci for Anthropometric Traits. *PLOS Genet* 2013; **9**: e1003500.

43	UK Biobank. Neale Lab. http://www.nealelab.is/uk-biobank (accessed Nov 11, 2024).

44	Yang M-L, Xu C, Gupte T, *et al.* Sex-specific genetic architecture of blood pressure. *Nat Med* 2024; **30**: 818–28.
